# Identification of novel microcephaly-linked protein ABBA that mediates cortical progenitor cell division and corticogenesis through NEDD9-RhoA

**DOI:** 10.1101/2022.03.28.22272122

**Authors:** Aurelie Carabalona, Henna Kallo, Maryanne Gonzalez, Liliia Andriichuk, Ellinoora Elomaa, Florence Molinari, Christiana Fragkou, Pekka Lappalainen, Marja W Wessels, Juha Saarikangas, Claudio Rivera

## Abstract

The cerebral cortex, which is responsible for higher cognitive functions, relies on the coordinated asymmetric division cycles of polarized radial glial progenitor cells for proper development. Defects in the mitotic process of neuronal stem cells have been linked to the underlying causes of microcephaly; however, the exact mechanisms involved are not fully understood. In this study, we present a new discovery regarding the role of the membrane-deforming cytoskeletal regulator protein called Abba (also known as MTSS1L/MTSS2) in cortical development. When Abba was absent in the developing brain, it led to a halt in radial glial cell proliferation, disorganized radial fibers, and abnormal migration of neuronal progenitors. During cell division, Abba localized to the cleavage furrow, where it recruited the scaffolding protein Nedd9, and positively influenced the activity of RhoA, a crucial regulator of cell division. Notably, we identified a variant of Abba (R671W) in a patient with microcephaly and intellectual disability, further highlighting its significance. The introduction of this mutant Abba protein in mice resulted in phenotypic similarities to the effects of Abba knockdown. Overall, these findings offer valuable mechanistic insights into the development of microcephaly and the cerebral cortex by identifying Abba as a novel regulator involved in ensuring the accurate progression of mitosis in neuronal progenitor cells.

## Introduction

Development of the central nervous system occurs by proliferation of neural progenitor cells, migration and differentiation of their progeny over substantial distance. These different steps require extensive membrane remodelling and cytoskeleton dynamics^1^. Neuronal migration involves the coordinated extension and adhesion of the leading process along the radial glial scaffold with the forward translocation of the nucleus, which requires regulation of centrosome and microtubule dynamics by different proteins^1–3^. However, little is known about the molecular mechanisms underlying membrane dynamics during neuronal migration and morphogenesis.

Cell membrane curvature is a micro-morphological change involved in many important cellular processes including endocytosis, exocytosis, and migration^4^. Recent studies demonstrated that members of an extended protein family, characterized by the presence of a membrane binding and deforming BAR (Bin–Amphiphysin–Rvs) domain function at the interface between the actin cytoskeleton and plasma membrane during the formation of membrane protrusions or invaginations^5–7^. These proteins have been shown to be important for proper neuronal morphogenesis^8^, and can either generate positive membrane curvature to facilitate the formation of plasma membrane invaginations (e.g. BAR, N-BAR and F-BAR domain proteins), or induce negative membrane curvature to promote the formation of plasma membrane protrusions (I-BAR and IF-BAR domain proteins)^9,10^. An important part of the membrane deforming mechanism include the interaction with phosphatidylinositol-4,5-biphosphate PI(4,5)P2-rich membranes through their BAR domain and also with actin cytoskeleton through Wasp Homology-2 domain (WH2).

Regulation of the actin cytoskeleton plays a crucial role in several developmentally important cellular processes including neuronal migration, axonal and dendritic extension and guidance, and dendritic spines formation^11^. We previously discovered that MIM (Missing-In-Metastasis; Mtss1), a member of the I-BAR protein family, has an important role in bending the dendritic plasma membrane in order to form dendritic spines and sculpt their morphology^12^. In a more recent study, a closely related protein Abba (Mtss1L) was found to be important for exercise induced increase synaptic potentiation in the hippocampus^13^. Although an important role of MIM and Abba in synapse formation and plasticity starts to emerge, information about the role of the early expression of this protein family during development is poor.

Mutations in BAR domain proteins are increasingly found in mental retardation disorders. For instance, altered expression and mutation of F-BAR family proteins, SrGAP2 & 3, are known to be associated to neurological disorders (mental retardation, the 3p syndrome, schizophrenia, early infantile epileptic encephalopathy and severe psychomotor delay^14–18)^. Furthermore, previous whole-exome sequencing of pre-screened multiplex consanguineous families identified *ABBA* as a likely candidate linked to neurogenic disorders^19^, suggesting that some BAR containing proteins might play important functions during brain development. Despite Abba’s potential role in neuronal development, very little is known about how this protein may lead to neurological pathology.

Regulation of the actin cytoskeleton and plasma membrane dynamics in glial cells of the developing brain has remained poorly understood. Interestingly, the I-BAR protein Abba is the only glia-enriched regulator that has been reported so far^20^.

We sought to determine the consequences of altered expression of *Abba* in radial glial progenitor (RGP) cells on cell cycle progression and neurogenesis and to test for subsequent neuronal migration. To address these issues, we used *in utero* electroporation to silence *Abba* with shRNA in embryonic mouse brains. Downregulation of Abba’s expression had a striking effect on RGP cells progression, resulting in a blocking of cytokinesis and a subsequent apoptosis. Biochemical analysis demonstrated that the cytokinesis block was likely due to an absence of interaction with the adhesion docking protein named Nedd9 (aka CAS-L or HEF1) that is well known to be a positive regulator of RhoA signaling in mitosis^21^. Using FRET based monitoring of RhoA activity, we also found a significant role of Abba on RhoA activity. Moreover, we observed a defect in neuronal migration associated with an increase in neuronal apoptosis. Importantly, we identified one patient harboring a missense variant in *ABBA* sequence and showing specific features such as: small head circumference, mental retardation, autism spectrum disorder (ASD) and craniofacial dysmorphism. We provide evidence that the (2011C>T(R671W)) patient mutation displays similar phenotypes and decreased Abba expression.

Overall, our data highlights the critical role of *Abba* in proper cortical development in neurogenesis and migration, and provides evidence that *ABBA* is a new causative gene for cortical malformation associated with ASD.

## Materials and Methods

### Plasmids construction

We utilized a single short hairpin RNA (shRNA) that targeted the coding sequence for Abba20. It is worth noting that the rat and mouse Abba sequences share a high degree of similarity, with a 95% homology. The specific region targeted by the shRNA displayed 100% homology. Both Abba and Nedd9 shRNA were designed using the siRNA Wizard Software (InvivoGen ©). As a negative control, we employed a non-targeting shRNA. These shRNAs were subcloned into an mU6pro vector^22^. Additionally, a pCAG-RFP vector was co-injected to facilitate the visualization of fluorescent cells. Abba mouse full length with silent mutations^20^, ABBA human full length and human ABBA-R671W carrying vectors were made by GeneCust© (France) and subcloned into the pCAGIG-IRES-GFP vector (Addgene, Cambridge, MA, USA).

### Bioinformatic methods

ABBA coding sequences were obtained from NCBI. The multiple sequence comparison was done by Log-Expectation (MUSCLE) method and visualized with DNASTAR MegAlign Pro. The impact of ABBA R671W on the structure and function of a human ABBA protein was predicted using PolyPhen-2 and HumVar model^23^.

### Human subjects

Families were identified through our clinical and research programs, personal communication, as well as the MatchMaker Exchange (MME) including GeneMatcher^24^ (http://www.genematcher.org). Informed consent for publication and analysis of photos, imaging and clinical data was obtained from the patients’ legal guardians. Brain magnetic resonance imaging (MRI) studies were performed on the subject and reviewed by the investigators in accordance with Department Ethical permission number (MEC-2012387) to conduct human exome studies.

### RhoA activity assay

Fluorescence from Raichu-Rac1 was imaged using a confocal microscope (LSM 710/Axio; Carl Zeiss) controlled by ZEN 2011 software (Carl Zeiss) and equipped with water immersion objective (63×/1.0 NA; Carl Zeiss). Transfected cells were illuminated with an argon laser (Lasos; Lasertechnik GmbH) at 458 nm (CFP and FRET) and 514 nm (YFP) using 0.5–10% of full laser power. Emission was collected at 461–519 nm (CFP) and 519–621 nm (FRET and YFP). The pinhole was fully opened. Scanning was performed in XY mode using 6× digital zoom that resulted in a pixel XY size of 68 × 68 nm. FRET signal was calculated as reported elsewhere23. In brief, using ImageJ 1.48v software images where background subtracted and FRET image generated using three images: MDonor (CFP excitation and CFP emission filters), MIndirectAcceptor (CFP excitation and YFP emission filters), and MDirectAcceptor (YFP excitation and YFP emission filters), using the following equation: FRET = (MIndirectAcceptor − MDonor × β − MDirectAcceptor × (γ − αβ))/(1 − βδ). Coefficients α, β, δ, and γ were obtained by independent control experiments analyzing cells expressing CFP or YFP as described previously25. Coefficients α, δ, and γ were determined in cells expressing only YFP (acceptor) and calculated according to equations: α = MDonor/MDirectAcceptor; γ = MIndirectAcceptor/MDirectAcceptor; δ = MDonor/MIndirectAcceptor. Coefficient β was determined in cells expressing only CFP (donor) and calculated according to the equation: β = MIndirectAcceptor/MDonor. After generation of FRET image, intensity of FRET signal was calculated using a mask obtained from a corresponding YFP image and covering neuronal cell bodies. The images were also analysed using Pix-Fret^26^.

### *In Utero* Electroporation

Animal experiments were performed in agreement with European directive 2010/63/UE and received approval N°: APAFIS#2797 from the French Ministry for Research and in agreement with the National Animal Experiment Board, Finland (license number ESAVI/18276/2018).Plasmids were transfected using intraventricular injection followed by *in utero* electroporation^27,28^. Timed pregnant C57BL6N mice (Janvier Labs; E14; E0 was defined as the day of confirmation of sperm-positive vaginal plug) received buprenorphine (Buprecare,0.03 mg/kg) and were anesthetized with sevoflurane/isoflurane (4%-4.5% induction, 2% -anaesthesia maintenance) 30 minutes later. For pain management, bupivacaine (2 mg/kg) was administered via a subcutaneous injection at the site of the future incision. Uterine horns were exposed, the lateral ventricle of each embryo was injected using pulled glass capillaries and a microinjector (Picospritzer II; General Valve Corporation, Fairfield, NJ, USA) with Fast Green (2 mg/ml; Sigma, St Louis, MO, USA) combined with the different DNA constructs at 1.5 μg/μl or with 1.5 μg/μl shRNA constructs together with 0.5 μg/μl pCAGGS-RFP and Anilline-GFP48. Plasmids were further electroporated by discharging a 4000 mF capacitor charged to 50V with a BTX ECM 830 electroporator (BTX Harvard Apparatus, Holliston, MA, USA). The voltage was discharged in five electrical pulses at 950ms intervals via 5mm tweezer-type electrodes laterally pinching the head of each embryo through the uterus. The mice were followed-up post-surgically and given buprenorphine (Buprecare 0.03 mg/kg) if needed. The number of animals has been calculated on the basis of the requirement for adequate numbers of brain slices and sections for sufficient imaging to provide statistically significant data on the effects of RNAi, small molecules, and other reagents used in the proposed analysis, plus controls.

### Organotypic cultures and Live Imaging

To assess the impact of altered ABBA expression in the RGP cells on neurogenesis and cell cycle progression, we utilized a combination of *in utero* electroporation and time-lapse imaging of organotypic cultures. The organotypic cultures were prepared as described^28^ with the following exceptions. After surrounding the brains (E17) with 4.5% UltraPure Low Melting Point Agarose (Invitrogen, 16520-100), 350 μm thick slices were cut with Vibratome 1000 Plus Sectioning System (No. 054018). Before placing the slices on a coated (83 µg/ml Laminin, Sigma L2020; 8,3 µg/ml Poly-L-Lysine, Sigma P4707) cell culture insert (Millicell, PICMORG50), sections were carefully cleaned from agarose to ensure adequate oxygenation. The inserts with slice cultures were transferred into a 6-well plate containing 1 ml of warmed slice culture medium. For live-imaging we used Andor Dragonfly 550 high speed Spinning Disc confocal equipped with Nikon Perfect Focus System, motorized XY stage, and environmental chamber. The system uses Fusion 2.0 software (version 2.1.0.48). The image acquisition was executed using Andor iXon 888 U3 EMCCD camera, 20x magnification with voxel size 0.65 × 0.65 × 0.9524 micron^3 (channel/excitation: RFP/561 nm, GFP/488 nm). Time lapse-imaging of organotypic cultures was started at E17+DIV0 with intervals of 10 minutes and total duration of 15 hours. The data was collected from 6 pups (2 scramble, 4 shRNA), 17 slices (5 scramble, 12 shRNA) and 24 areas (9 scramble, 15 shRNA; some slices were recorded from two different areas). For quantification of Anillin-GFP^48^ positive cells, Z-projection images of individual timepoints were made and positive cells tracked over time using the plugin TrakMate in ImageJ (NIH, Bethesda, MD).

### Immunostaining of Brain Slices

Mice brains were fixed (E17) in Antigen Fix (Diapath), Brain slices were sectioned coronally (80μm) on a vibratome (Leica microsystems). Brain slices were washed with PBS (Phosphate-buffered saline pH 7.4) and stained in PBS 0.3% Triton X-100 supplemented with 5% of donkey serum. Primary antibodies were incubated overnight at 4°C, sections were then washed with PBS and incubated in secondary antibodies for 2 hours at room temperature. Antibodies used in this study were: Abba (homemade), Vimentin (Millipore, MAB3400), Ki67 (Millipore, AB9260), phospho-histone H3 (Abcam, ab14955), Pax6 (biolegend, PRB-278P).

### Cells culture and transfection

Rat C6 glioma cells were cultured at 37°C under a humidified atmosphere with 5% CO2 with a complete medium DMEM supplemented with 10% foetal bovine serum (FBS, Sigma) and 100 units/mL antibiotics/antimycotics. Cells were transfected using the Neon® Transfection System (Life Technologies) according to the manufacturer’s protocol. Briefly, cells were trypsinized and counted with the cell counter SCEPTER (Millipore). Electroporation was performed with 500 000 cells and a total amount of 5 µg of DNA containing a reporter plasmid encoding RFP (1:5) and shRNAs or mismatch constructs, with the following configuration: 1860 V, 1 pulse, 20 ms. Cells were then cultured on a 6 wells plate during 2 days before RNA extraction.

### Immunocytochemistry

HEK293T cells were fixed in Antigen Fix (Diapath), for 15 min and blocked at room temperature for 1 h with 5% normal goat serum,0.3% Triton X-100 in PBS and incubated overnight at 4°C with a set of antibodies against Abba (homemade), Phalloidin (Thermo Fisher, A12379), Nedd9 (Abcam, ab18056), Hoechst (Thermo Fisher, 62249). Cells were then washed with PBS and incubated in secondary antibodies for 2 hours at room temperature. The definition of cortical regions was guided by stainings against Cux1 (santa cruz, sc-13024) and Ctip2 (Abcam, ab18465) .

### RT-PCR and qPCR

Total RNA was isolated from C6 using RNeasy Plus Mini kit and cDNA was synthesized using the Quantitect Reverse Transcription kit, according to the manufacturer’s protocol (QIAGEN). Quantitative PCR (qPCR) was performed on a Light cycler 480 using SYBR-Green chemistry (Roche) and specific primers for Abba (F: CGAGACTCGCTGCAGTATTCC and R: CCATTCACAGAGTAGCAGTCG, 113 bp), Nedd9 (Qiagen, QT01610385) and CyclophilinA (Qiagen, QT00177394) as a control probe. qPCR was performed with 5 µL of diluted cDNA template, specific primers (0.6 µM) and SYBR Green I Master Mix (7.5 µL) at a final volume of 15 µL. Each reaction was performed at an annealing temperature of 60°C and for 50 cycles. Reactions were performed in duplicate and melting-curve analysis was performed to assess the specificity of each amplification. A standard curve was performed for each gene with a control cDNA diluted at different concentrations. Relative expression was assessed with the calculated concentration in respect to the standard. All experiments were performed in triplicate.

### Microscopy and Image Analysis

All images were collected with an IX80 laser scanning confocal microscope (LSM 710/Axio; Carl Zeiss) controlled by ZEN 2011 software (Carl Zeiss)). Cells & Brain sections were imaged using a 60x 1.42 N.A. oil objective or a 10x 0.40 N.A. air objective. All images were analyzed using ImageJ (NIH, Bethesda, MD). To quantify the total number of C6 cells alive and in mitosis on 12mm diameter coverslips 2 & 3 days after Abba shRNA & control transfections, we have arbitrary designed 4 identical regions comprised of 9 squares using ZEN software acquisition system and count manually the total number of alive and mitotic cells in each condition. All cell quantifications in brain slices were initially normalized to electroporation efficacy. For analysis of morphological changes in vimentin staining images were analysed using the directionality plugin in ImageJ and the values corresponding to dispersion extracted.

### Statistical data

Statistical analyses were performed with Prism (GraphPad Software, La Jolla, CA, USA). A two-sample Student’s t-test was used to compare means of two independent groups if the distribution of the data was normal. If the values come from a Gaussian distribution (D’agostino-Pearson omnibus normality test) the parametric unpaired t-test with Welch’s correction was used. But, when the normality test failed, the non-parametric Mann–Whitney test was used. Significance was accepted at the level of p < 0.05.

No statistical methods were used to predetermine sample sizes, but our sample sizes are similar to those generally employed in the field. No randomization was used to collect all the data, but they were quantified blindly.

### Yeast two hybrid screen

Yeast two hybrid screening was performed by Hybrigenics Services (Paris, France). The Abba coding sequence containing amino acids (aa) 1 715 was cloned into pB27 (N-LexA-bait-C fusion) and pB66 (N-GAL4-bait-C fusion) and sequence verified. These constructs were used as baits to screen the mouse embryo brain RP2 prey library. Total of 91 million (pB27) and 31 (pB66) million interactions were analyzed and the detected interactions were assigned with a statistical confidence score, the Predicted Biological Score (PBS) (www.hybrigenics-services.com).

### Co-immunoprecipitation

C6 cells or cortical tissue were washed with ice cold PBS and lysed using lysis buffer (20 mM Tris-HCl [pH 8,0], 137 mM NaCl, 10% glycerol, 1% Triton X-100, 2 mM EDTA) with 50 µg/ml PMSF (Roche) and a protease inhibitor cocktail (Roche). Cell lysate was agitated for 30 min at 4°C and centrifuged at 12000 rpm for 20 min at 4°C. The supernatant was collected and protein concentrations were determined with a Pierce^TM^ BCA Protein Assay kit, according to the manufacturer’s protocol (Thermo Scientific). Before Co-IP, lysates were precleared with washed Pierce Protein A/G Magnetic Beads (Thermo Scientific) overnight in gentle shaking at 4°C, to remove nonspecific binding. Beads used for Co-IP were washed and pre-blocked with 3% BSA in PBS overnight in gentle shaking at 4 °C. Washing steps were done using a wash buffer (1% BSA, 150 mM NaCl, 0,1% Tween-20 in TBS).

For Co-IP, 1000 µg of protein from precleared lysate was incubated with 20 µl of specific primary antibodies (Abba: homemade; Nedd9: Abcam, ab18056) overnight in gentle shaking at 4°C. Negative controls were incubated without primary antibodies under the same conditions. Immunocomplexes were pulled down by incubating the antigen-antibody mixture with pre-blocked beads for 1 h in gentle shaking at room temperature. Beads were washed 3 times with a wash buffer and once with purified water before elution. Bound proteins were eluted from the beads by incubating them in 1x Laemmli buffer containing 150 mM NaCl for 10 min at room temperature.

Original C6 cell lysate and immunoprecipitated proteins were resolved by SDS-PAGE (4-20% Mini-PROTEAN TGX Gel; Bio-Rad) and transferred using Trans-Blot Turbo Mini PVDF Transfer Packs and Transfer System (Bio-Rad). Membranes were blocked with TBS containing 0,1% Tween-20 and 5% milk and blotted with primary antibodies overnight in shaking at 4°C. HRP-conjugated secondary antibodies used in this study were Rabbit anti-mouse IgG and Goat anti-rabbit IgG (Invitrogen). Protein bands were visualized using the ChemiDoc^TM^ MP imaging system (Bio-Rad).

### Flow-cytometry

To measure cell cycle length, 4×105 cells were seeded in 6-well plates, transfected and cultured for 24h. Plasmid transfection was performed with jetPEI by following the Batch HTS protocol suggested in the data sheet of the product. Briefly, transfection complexes, which consist of both DNA and jetPEI reagent diluted in NaCl, are mixed with the suspension of cells obtained after trypsinization and distributed in the wells for culture over 24h. Then, cell cycle synchronization was induced by 24h serum starvation and re-started using fetal calf serum supplemented DMEM. The cells were detached with trypsin+EDTA and fixed with 3.7% formaldehyde after 24 hours in culture. 5×105 fixed cells from each stop point were stained in 200ul of Hoechst 33342 (1:2000 dilution) for 30min, washed with PBS 1X and diluted in a flow buffer. Acquisition of 100 000 total events was performed in NovoCyte Quanteon 4025 and the analysis performed in (FlowJo v10.8 or NovoExpress 1.6.1) software. Graphs and statistical analysis were performed in GraphPad Prism 5.

## Results

### *Abba* knockdown disrupted radial glia morphology

Previous studies of the expression pattern of *Abba* in the developing brain has disclosed a conspicuous expression in radial glia. In order to investigate the function of *Abba* in the developing brain, we used a mouse *in utero* RNAi approach to knockdown *Abba* at E14. For this purpose, we opted for shRNA knockdown using a previously validated targeting sequence^20^. Revalidation of the knockdown efficiency of this target with three different Abba-shRNA (Abba-shRNA1-3) in rat C6 glioma cells showed a 70– 80% reduction of Abba mRNA and protein levels with Abba-shRNA3 (Supplemental Figure 2, Abba-shRNA3).

To determine whether knockdown of Abba expression alters neuronal migration, Abba- shRNA3 combined with a red fluorescent protein (RFP) construct was introduced into neural progenitor cells of mouse neocortex by *in utero* electroporation at E14 and evaluated at E16-18 (Figure 1A). Electroporation of E14 targets the progenitors that will migrate and populate cortical layers II/III neurons. In E17 brain sections, we observed that some neurons previously electroporated 3 days earlier with the scrambled construct reached the cortical plate as expected (Figure 1B). However, *in utero* expression of Abba-shRNA3 induced a significant arrest of cells within the ventricular zone (VZ) (Figure 1A-C; scramble: 24,3 ± 6.4%; n=9, Abba-shRNA3: 63,7 ± 2,9%; p<0.0001; n=8). Surprisingly, we also observed a striking decrease in the total number of RFP positive cells in brain sections electroporated with Abba-shRNA3 at E17 and E18 (Figure 1D-E; scramble: 969,2±109.0 cells/mm^2^; n=9, Abba-shRNA3: 311,3±28,61 cells/mm^2^; p<0.0001; n=8). These results indicate an important role of Abba in the proliferation and migration of cortical neuronal progenitors.

**Figure 1.**
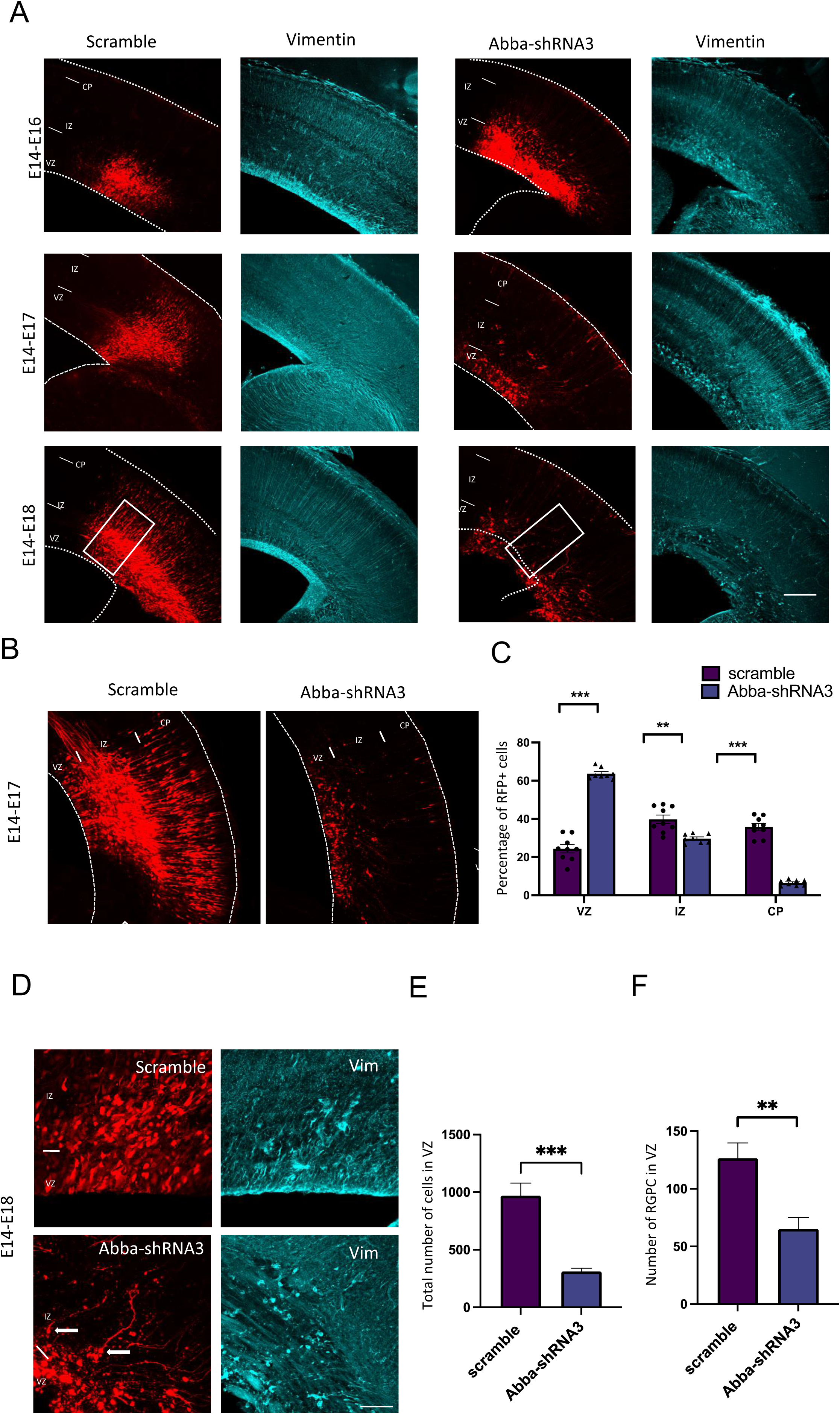
*In utero* knockdown of *Abba* expression is linked to radial glial disruption and alters neuronal migration. **A.** Representative coronal sections of E16-E18 mice brains electroporated at E14 with either scramble or Abba- shRNA3 and immunostained for vimentin (in blue). In Abba-shRNA3 brain sections, vimentin staining revealed a marked disruption of radial glial apical fibers. **B.** Representative neocortical coronal sections showing migration of transfected cells 3 days after electroporation at E14 with either scramble or Abba-shRNA. **C.** Quantification of RFP-positive cell distribution in the cortex at E17 (VZ/SVZ: scramble 24.33±2.13%, Abba-shRNA3 63.71±1.03%; IZ: scramble 39.80±2.22%, Abba-shRNA3 29.71±0.85%; CP: scramble 35.86±1.73%; Abba-shRNA3 6.58±0.53%; n=9 scramble and 8 Abba-shRNA3; (VZ/SVZ: p=<0.0001: IZ: p=0.0002; CP: p=<0.0001) showing a significant increase of Abba knockdown cells in SVZ/lower IZ. **D.** Higher magnifications of regions indicated by a square in C. **E.** Total number of RFP positive cells in mouse neocortex at E17 (scramble 969.2±109.0%; Abba-shRNA3 311.3±28.61%; n=9 scramble and 8 Abba-shRNA3) showing a striking reduction of RFP positive cells in Abba knockdown electroporated brains. Error bars represent mean ± s.d. **P < 0.002, ***P < 0.001, Scale bars 100μm (A). **F.** Quantification of radial glial progenitor cells at the VZ (scramble 126.50±13.36%, Abba-shRNA3 65.14±9.91%). mean ±SD. VZ: ventricular Zone, IZ: Intermediate Zone, CP: Cortical Plate. Error bars represent mean ± s.d. **P < 0.002. Scale bars represent100μm (A), 50μm (B).

We also evaluated the impact of *Abba* knockdown on radial glial morphology by imaging vimentin-stained *Abba* knockdown cells at 3 days post-electroporation. All E17 *Abba* knockdown brains, but not in age-matched controls, displayed disruption of radial glial fibers in the VZ/SVZ with increased dispersion in Abba knockdown expressing (Figure 1D and F; Supplemental Figure 1; Scramble 14.27.4±6 % and Abba-shRNA3 40.8 ±19, n=20 P=0.0078). Control vimentin-positive cells displayed the characteristic features and polarized morphology of radial glia with processes extending from the VZ to the pial surface. In contrast, in *Abba* knockdown brains, vimentin-positive cells exhibited misoriented apical processes, detached end feet in the ventricular surface (Figure 1D) and decreased number of radial glial progenitor cells in the VZ (Figure 1F; scramble: 126,5 ± 13,4 cells, n=8; Abba-shRNA3: 65,1 ± 26,2 cells, n=7; p=0.0033). All together these data show that the loss of Abba can disrupt radial glial cells morphology thus affecting their progenitor function.

These data suggest that *Abba* is required for radial glial organization, neuronal migration, regulation of neuronal proliferation and cell survival. Importantly, these results are in accord with the reported association of *ABBA* with developmental neurological diseases including intellectual disability and microcephaly^19^.

### ABBA is required for cell cycle progression of radial glia progenitors

Development of the cerebral cortex occurs through a series of stages, beginning with RGPs. These stem cells exhibit an unusual form of cell-cycle-dependent nuclear oscillation between the apical and the basal regions of the ventricular zone, known as interkinetic nuclear migration (INM). RGP cells undergo mitosis only when they reach the SVZ^30,31^. It has been shown that inhibition of INM disrupts RGP cell cycle progression^32–34^.

To test the effect of altered radial glial cell morphology on cell cycle progression, we have quantified the nuclear distance from the VS for RGP cells after electroporation of scrambled or Abba RNAi (Figure 2A). We observed an accumulation of Abba- shRNA3 RGP cells close to the ventricular surface (0-10 mm; scramble: 30.2 ± 3.2%, n=7; Abba-shRNA3: 38.3 ± 2.3%, n=7; p= 0,000156) which can correspond to mitotic cell, but also further away (>30 mm; scramble 12,0 ± 3.3%, n=7; Abba-shRNA3: 21,9 ± 2.4%, n=7; p= 0,000023) which support our previous observations on cells detaching from the VS (Figure 1D and F). Furthermore, we observed that Abba-shRNA3 decrease the percentage of the RGP cells marker Pax6 (Figure 2B and C; scramble: 31,9 ± 0,6%, n=8; Abba-shRNA3: 21,1 ± 0,6%, n=5; p=<0.0001). We have then tested for cell cycle effects, and found a decrease on the percentage of Abba shRNA3-expressing RGP cells positive for the cell cycle marker Ki67 (Figure 2D and E; scramble: 46,5 ± 0,8%, n=7; Abba-shRNA3: 23,7 ± 1,369%, n=5; p= <0.0001) and also for the late G2/M phases marker phospho-histone 3 (PH3, Figure 2 F and G; scramble: 4,5 ± 0,1764%, n=9; Abba-shRNA3: 1,264 ± 0,5191%, n=5; p= <0.0001). More detailed examination of the impact of Abba-shRNA3 on cell cycle progression using flow cytometry showed an accumulation in S phase. Interestingly, these effect was rescued by overexpression of Abba-FL (Supplemental Figure 3; G1: scramble: 45,2 ± 1,4 %, Abba-shRNA3: 37,4 ± 1,0 %, Abba-shRNA3+Abba-FL: 42,8 ± 1.3 %; S: scramble: 34,3 ± 2,8 %, Abba- shRNA3: 48,2 ± 2.7 %, Abba-shRNA3+Abba-FL: 38,9 ± 3.4; G2: scramble: 17,1 ± 2,1 %, Abba-shRNA3: 14,4 ± 2,7%, Abba-shRNA3+Abba-FL: 15,9 ± 1,9 %; n=10; p=<0.001). These results indicate that Abba plays a major role in coordinating cell cycle progression in radial glia.

**Figure 2.**
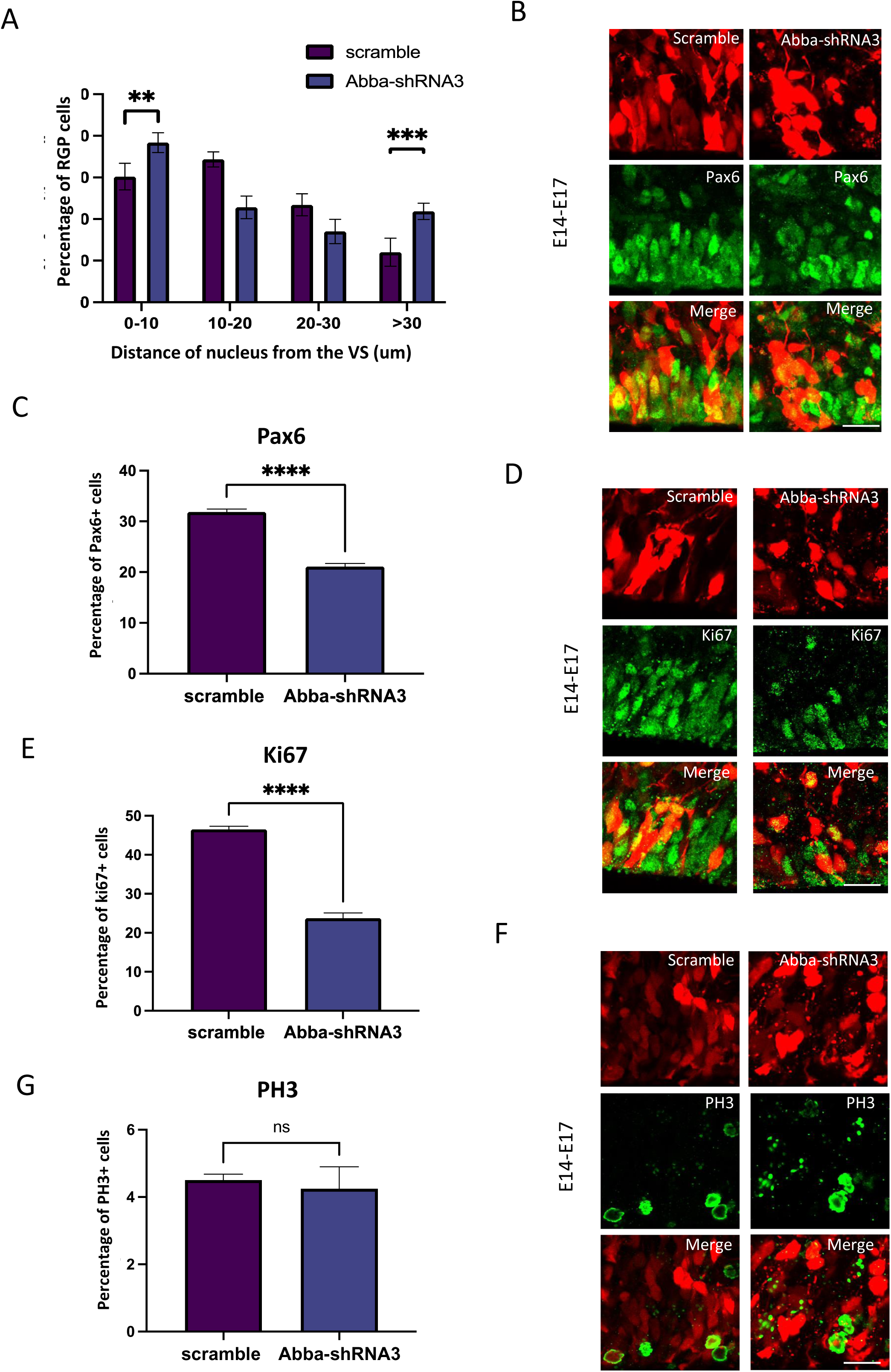
*Abba* down-regulation induced inhibition of basal nuclear migration in RGP cells blocks cell cycle progression. **A.** Quantification of the nuclear distance from the VS for RGP cells after electroporation of scrambled or Abba-shRNA3. Nuclear distribution of RFP RGP cells was significantly altered after electroporation of Abba- shRNA3 (0–10: scramble, 30.20 ± 1.21%; Abba-shRNA3, 38.34 ± 0.89%; 10–20: scramble, 34.35 ± 0,69%; Abba-shRNA3, 22.80 ± 1.03%; 20–30: scramble, 23.42 ± 1.00%; Abba-shRNA3, 17.00 ± 1.10%; >30: scramble, 12.04 ± 1.27%; Abba-shRNA3, 21.85 ± 0.74%; (0–10: P = 0,000156; 10-20: P=<0.000001; 20-30:P= 0,001030; >30:P= 0,000023). **B-C.** E14 mice brains were subjected to *in utero* electroporation with either scramble or Abba- shRNA3. Brains were then fixed at E17 and stained with the RGP cell marker Pax6. We observe an important decrease of Pax6 RGP cells expressing Abba-shRNA3(scramble, 31.87 ± 0.57%; Abba-shRNA3, 21.08 ± 0.63%; n = 8 scramble; 5 Abba-shRNA3). **D-G.** E14 mice brains were subjected to *in utero* electroporation with either scramble or Abba- shRNA3. Brains were then fixed at E17 and stained with the cell cycle marker Ki67 (D)and PH3 (F). We observe a striking decrease in the percent of cycling (E: Ki67; scramble, 46.54 ± 0.79%, n=7; Abba-shRNA3, 23.71 ± 1.37%, n=5 Abba-shRNA3)) and but not in mitotic (G:PH3; scramble, 4.50 ± 0.17%; Abba-shRNA3, 4.25 ± 0.65%) cells with low expression of Abba. G. Flow cytometry analysis of cells electroporated with Abba-shRNA3 show accumulation in S-phase and recovery after additional expression of Abba-FL (**G1**: scramble 45,2 ± 1,4 %, Abba-shRNA3 37,4 ± 1,0 %, Abba-shRNA3+Abba-FL 42,8 ± 1.3 %; **S**: scramble 34,3 ± 2,8 %, Abba-shRNA3 48,2 ± 2.7 %, Abba-shRNA3+Abba-FL 38,9 ± 3.4; **G2**: scramble 17,1 ± 2,1 %, Abba-shRNA3 14,4 ± 2,7%, Abba- shRNA3+Abba-FL 15,9 ± 1,9 %; n = 10). Error bars represent mean ± s.d. **P < 0.002, ***P < 0.001. Scale bar represents 20μm.

### Abba localizes to the cleavage furrow and is important for cytokinesis

To further characterize the role of Abba in mitotic progression, we synchronized C6 rat glioma cell line and stained with Abba antibody. We found that Abba localized to the putative apical membrane initiation site and cytokinetic bridge, proximal to the contractile actin ring during late telophase and cytokinesis (Figure 3A and B respectively). The actomyosin contractile ring is formed at the plasma membrane in response to accumulation of phosphoinositide 4,5 bisphosphate (PI(4,5)P2), to generate constricting force to separate the two daughter cells^35^. Abba is known to interact with PI(4,5)P2 via its membrane deforming I-BAR domain^20^ that has the capacity to deform and recognise the membrane curvature.

**Figure 3.**
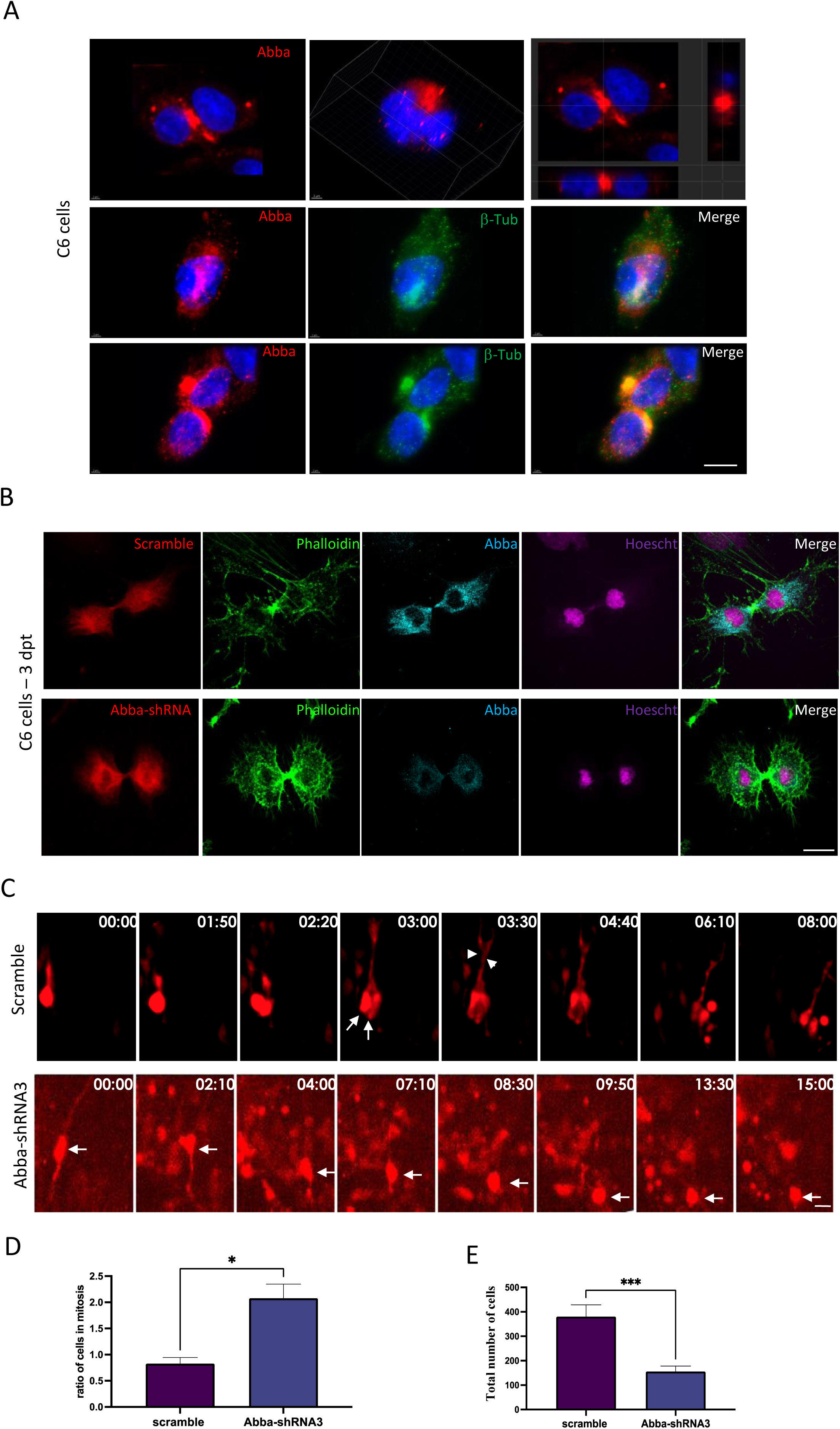
*Abba* down-regulation blocks cytokinesis. **A.** Cellular distribution of Abba in C6 cells during cell division. Upper panel shows two example immunofluorescent microscopy images of Abba (red). Left most figure shows the projection images at the cross line of the image. Mid and lower panels show the distribution of Abba at different cell division stages in relation to β-tubulin (green) aggregation **B.** Immunofluorescence microscopy images from C6 cells 72 hours after transfection with Abba-shRNA3 shows an absence of Abba expression at cytokinesis. **C.** Brain slices prepared from in utero electroporated animals with either Abba-shRNA or scramble were cultured at E17 and subjected to live imaging for a duration of 15-20 hours. The length of the time lapse was adjusted as necessary to capture significant events during interkinetic nuclear migration (INM). In the case of the scramble group, an RGP cell underwent a single mitotic event at the ventricular surface of the brain slice, observed at timepoints 1:50 and 2:20. At timepoint 3:00, two cells can be observed (indicated by arrows), along with the presence of basal fibers (indicated by arrowheads). Conversely, in the Abba knockdown group, the nucleus of the RGP (indicated by an arrow) initially underwent apical INM and subsequently divided at 8:30. Remarkably, the resulting daughter cells remained at the ventricular surface for at least 7 hours, suggesting that the absence of Abba hinders cells from exiting the mitotic state. **D**. Abba-shRNA3-expressing cells show increase in cytokinesis (scramble 0,8285 ± 0,1161%, n=12; Abba-shRNA3 2,078 ± 0,2682%, n=12) **E.** Quantification of the total number of RFP positive cells in the neocortex illustrating the impact of cytokinesis block on cell survival. We observed a lower total number of cells 4 days after transfection with Abba-shRNA3 compared with control scramble (scramble 380,3± 48,16%; n=12, Abba-shRNA3 155,8± 22,32%; p= <0.0001; n=12). Error bars represent mean ± s.d. *P < 0.03, **P < 0.002, ***P < 0.001. Scale bars 50μm (A), 20μm (D).

To determine the impact of the loss of Abba on cytokinesis, we quantified the number of C6 cells in cytokinesis 3 days after transfection with Abba-shRNA3 or scramble shRNA. Importantly we found a two-and-a-half-fold increase in the percentage of Abba-shRNA3-expressing cells in cytokinesis (Figure 3B and D; scramble 0,8285 ± 0,1161%, n=12; Abba-shRNA3 2,078 ± 0,2682%, n=12; p=0,0358), suggesting a prominent role of Abba in cytokinesis. To validate this *in vivo*, we performed live imaging in cortical organotypic slices from animals that were *in utero* electroporated with either Abba-shRNA3 or scramble vectors. Analysis of live imaging recording showed a strong mitotic block at the ventricular surface (Figure 3C) as well as significant accumulation of a marker of mitotic progression Anillin-GFP in RFP cells during the recording period (Supplementary Figure 6, Scramble: 100.2 ±15 % and Abba-shRNA3: 152.6 22, n=20 P=0.036). We then examined the impact of cytokinesis block on cell survival and found a lower total number of cells 4 days after transfection with Abba-shRNA3 compared with control scramble (Figure 3E; scramble: 380,3 ± 48,16%, n=12; Abba-shRNA3: 155,8 ± 22,32%, n=12; p=<0.0001). Taken together, these data provide evidence that Abba is required for the completion of mitosis, and its absence results in a reduced number of cell progeny.

### Abba recruits Nedd9 to the cytokinetic bridge and is required for RhoA activation

In earlier work we have shown that ABBA is mainly expressed through E10.5-12.5 in the floorplate structure formed by radial glia^20^. To investigate the mechanism by which Abba operates in mitosis, we performed a yeast-two-hybrid (Y2H) screen in E10.5-E12.5 mouse brain embryo library using full length mouse Abba as a bait. The resulting analysis identified three high (PBS: B) / good (PBS: C) confidence interactors: the E3 ubiquitin ligase Beta-Trcp2 (PBS: B), scaffolding protein Nedd9 (PBS: C), the transcription factor Otx2 (PBS: C) (Fig 4A; Supplemental table 2). Among these candidate proteins, Nedd9 (aka HEF-1 or Cas-L) caught our interest as it was previously reported in breast cancer cells to localize to the cleavage furrow during cytokinesis where it activates RhoA, as well as abnormal expression of Nedd9 results in cytokinesis defect^21,36,37^ (Figure 4B).

**Figure 4.**
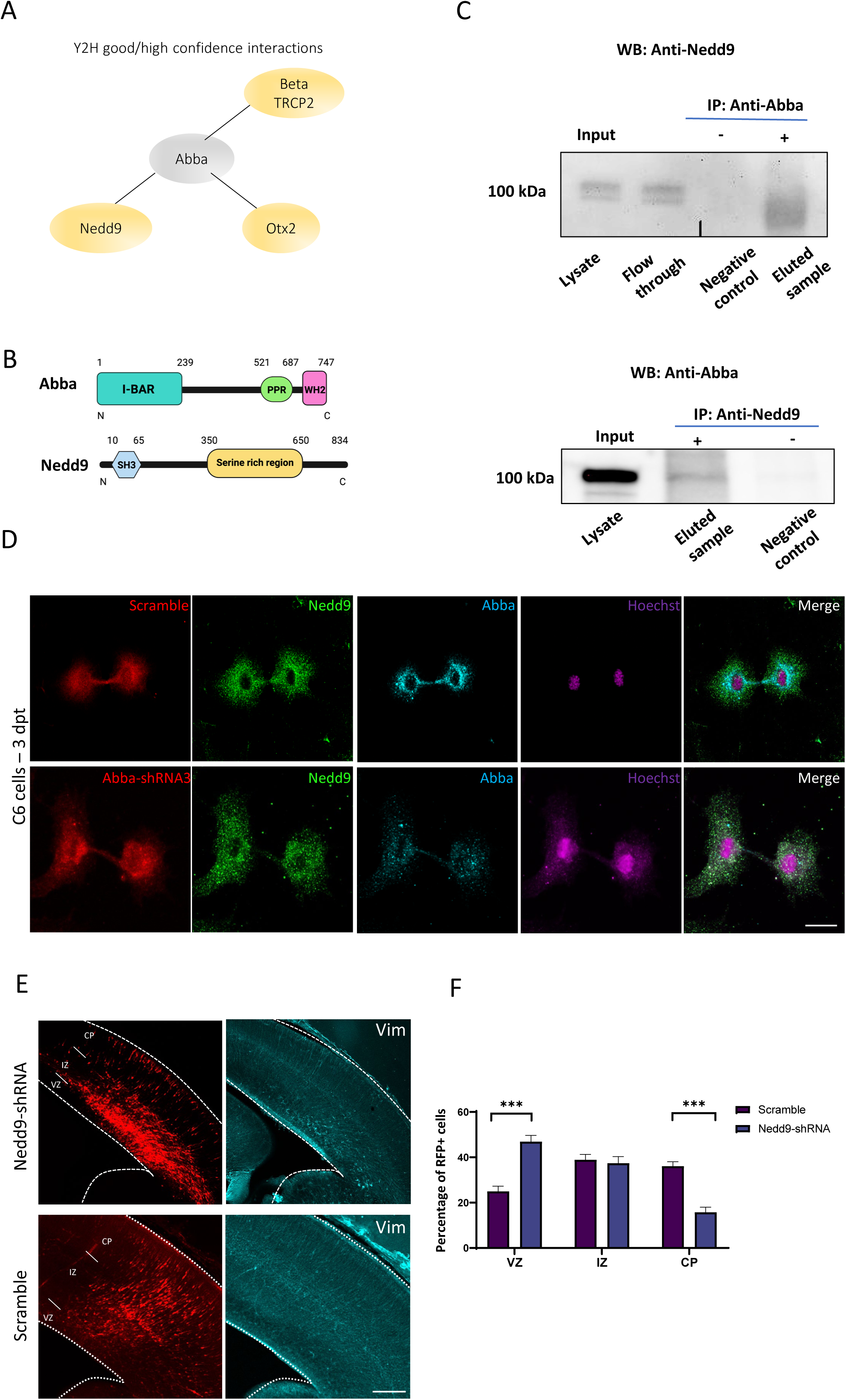
Role of Abba-Nedd9 complex in RGP cells division. **A.** Good and high confidence interactions detected in yeast two-hybrid screen performed in mouse brain embryo library using full length mouse Abba as a bait. **B.** Schematic representation of the Abba and Nedd9 proteins. **C.** E18 cortical homogenates were subjected to a direct pulldown assay with endogenous Abba and Nedd9. Nedd9 co-immunoprecipitated with Abba, but not in control pulldowns showing the specificity of the interaction. **D**. immunocytochemistry on C6 cells 72 hours after transfection with Abba-shRNA3 shows that lack of Abba did not decrease Nedd9 expression (green) but rather its localization at the cleavage furrow/cytokinetic bridge stage. Scale bars 20μm. **E.** Representative coronal sections of E17 mice brains transfected at E14 with Nedd9-shRNA and immunostained for vimentin (in blue). Vimentin staining revealed a marked disruption of radial glial apical fibers. **F.** Quantification of RFP-positive cell distribution in the cortex at E17 (VZ/SVZ: scramble 25 ± 2.30%, Nedd9-shRNA 46.92±2.78%; IZ: scramble 38.94±2.32%, Nedd9-shRNA 37.37±2.96%; CP: scramble 36.06±1.95%; Nedd9-shRNA 15.71±2.25%; n=8 scramble and 7 Nedd9-shRNA; (VZ/SVZ: p=<0.0001: IZ: p=0.0002; CP: p=<0.0001) showing a significant increase of Nedd9 knockdown cells in SVZ/lower IZ. Scale bars 100μm (E), 50μm (D).

We first tested whether Abba and Nedd9 physically interact, as suggested by the Y2H screen. To this end, we performed immunoprecipitation experiments from embryonic day 18 cortical tissue as well as C6 cells using anti-Nedd9 as well as anti-Abba antibodies. The precipitated Nedd9-Abba complex was separated on SDS-page and followed by western blot with anti-Abba and anti-Nedd9 antibodies respectively. Indeed, Abba precipitated with Nedd9 in both directions, but not using control beads from brain cortex homogenates (Figure 4C) and C6 cells (Supplemental figure 4). Nedd9 stability is not affected by the availability of Abba as western blots show no changes in Nedd9 expression levels in Abba-shRNA3 expressing cells (data not shown). Interestingly however, when we analyzed the localization of Nedd9, we found that Abba was required for targeting Nedd9 to the cleavage furrow/cytokinetic bridge in C6 cells 72 hours post transfection (Figure 4D). These data suggest that Abba recruits Nedd9 to the cleavage furrow during RGP cells division, which fits to its role in sensing curved PI(4,5)P2 rich membranes. Nedd9 plays a role in RGP cytokinesis *in vivo*^21,36,37^. To knockdown Nedd9, we used shRNA targeting Nedd9 which reduced mRNA levels by 75% in C6 cells (Supplemental Figure 4A). We used a previously characterised Nedd9-shRNA to determine whether knockdown of Nedd9 expression alters RGP cells morphology and function. To this end, Nedd9-shRNA combined with a red fluorescent protein (RFP) construct was introduced into neural progenitor cells of mouse neocortex by *in utero* electroporation at E14 and stained with the radial glial cell marker, vimentin. Interestingly, similarly to Abba knockdown, Nedd9 downregulation resulted in disruption of INM as a larger portion of progenitors accumulated close to the ventricular surface (Figure 4E-F; scramble: 25 ± 2.30% n=8, Nedd9-shRNA: 46.92±2.78%; n=7; p=<0.0001). Using the late G2/M phases marker, PH3, we observed similar results as with Abba shRNA and point mutation no difference in number of PH3 positive cells (Supplemental figure 4C; scramble: 4.50 ± 0.18%, n=9; Nedd9-shRNA: 5.07 ± 0.35%, n=6, P=0,1135). The role of Nedd9 in cell cycle progression is consistent with previous published results^21,36,37^ and provide evidence that both Abba and Nedd9 function to ensure correct mitotic completion of RGP cells.

Finally, to determine whether Abba is important for Nedd9 activity and how Abba and Nedd9 converge to regulate the cell-cycle, we assessed the activity of RhoA, a downstream signalling molecule of Nedd9^38^ which is important for cytokinetic actomyosin ring assembly and dynamics^38–40^. To determine RhoA activity we used a GFP-based FRET construct for RhoA designated ‘‘Ras and interacting protein chimeric unit’’ Raichu-RhoA. In order to test the efficacy of the assay we used Calpeptin, a RhoA activator which, as expected, increased RhoA activity (% change compared to Raichu-RhoA: Calpeptin 132,2 ± 3,2 n=33; p=0.002) confirming the functionality of our assay (Figure 5). Importantly, when Abba was silenced by shRNA there was a significant 31,5% decrease in Raichu-RhoA activity as compared to the control shRNA treated cells (Figure 5B; scramble: 103,1.03 ± 5.3; n=102, Abba-shRNA3: 68,5 ± 2,3; n=46; p<0.0001). In addition, this effect was rescued by the co-expression of shRNA insensitive plasmid carrying the full-length sequence of Abba (Abba-shRNA + Abba-FL 139,8 ± 19,6 n=26; p<0.0001 compared to Abba-shRNA3). Abba-FL did not change RhoA activity significantly compare to scramble expressing cells (Abba-FL 105,2 ± 16,0; n=37). Similar results were found when quantifying Raichu-RhoA activity in regions corresponding to the cleavage furrow (Figure 5 C; scramble: 76,3 ± 27,6; n=18, Abba- shRNA3: 44,8 ± 17,2; n=15, Abba-shRNA + ABBA-FL: 102.,5 ± 49,7 n=23, Abba-FL: 81,6 ± 38,7,0; n=22; p<0.0001, Abba-R671W: 135,7 ± 62,4 n=15). These data provide support for the function of Abba in spatial assembly of cytokinetic signaling through the Nedd9-RhoA axis.

**Figure 5.**
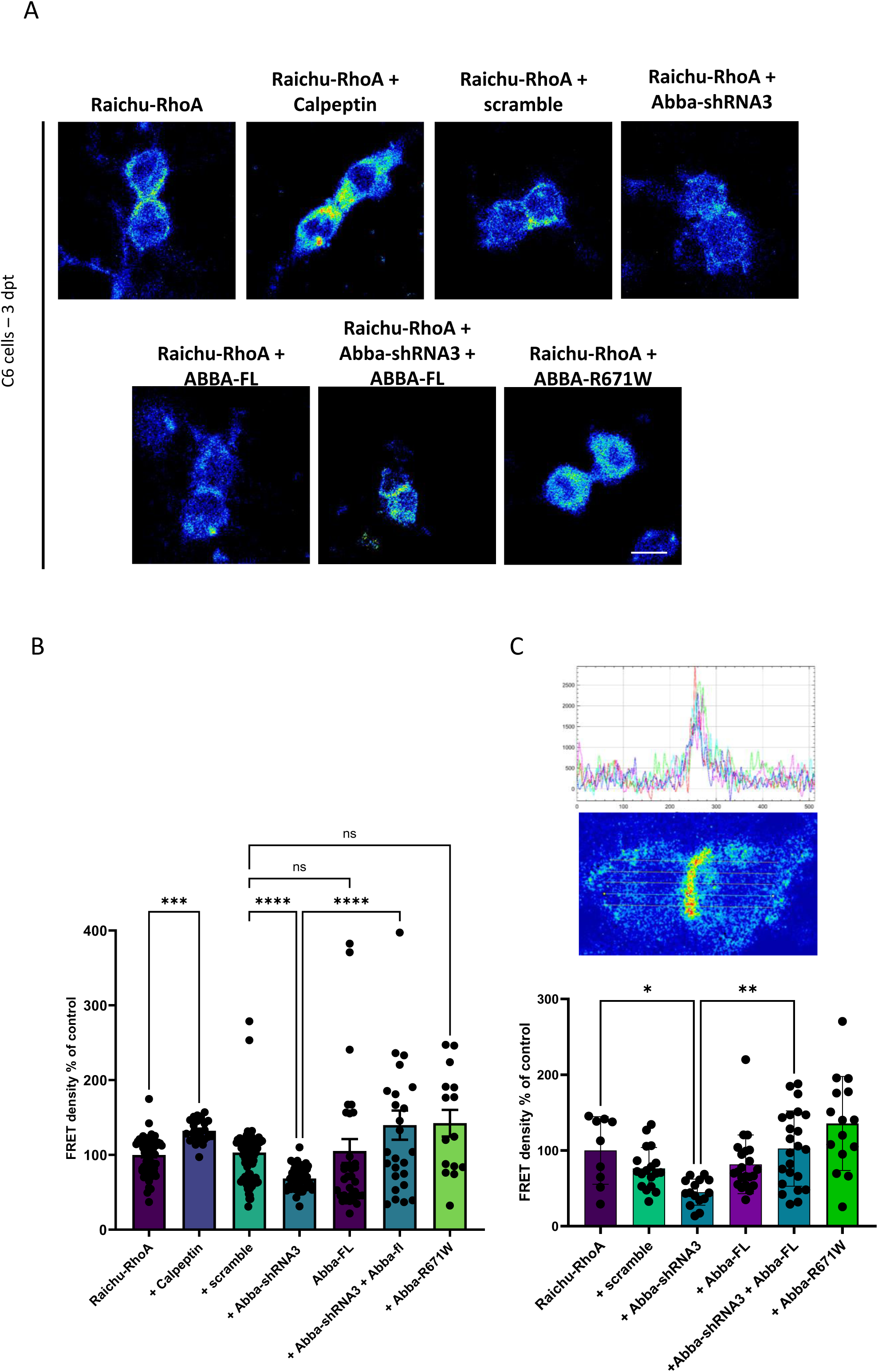
Abba downregulation decreases RhoA activity. **A.** Confocal images were captured of C6 cells 72 hours post-transfection with the Raichu-RhoA vector for monitoring RhoA activity. Six experimental groups were examined: transfected with scramble control, Abba- shRNA3, Abba-fl, rescue of Abba-shRNA3 with Abba-fl and expression of the mutant Abba-R671W. In addition to testing the sensitivity of the assay to RhoA activation by calpeptin. **B.** Quantification of FRET density (Raichu-RhoA activity) showed a decreased of Raichu-RhoA activity in C6 cells transfected with Abba-shRNA3 as compared to transfected with the scramble (scramble: 103,1.03 ± 5.3; n=102, Abba-shRNA3: 68,5 ± 2,3; n=46; p<0.0001, Abba-shRNA + ABBA-FL: 139,8 ± 19,6 n=26, Abba-FL: 105,2 ± 16,0; n=37, Abba-R671W: 142 ± 17,9 n=15). C. Quantification of the Raichu-RhoA activity in the furrow region show similar effects. **C**. upper panels show FRET image of a dividing cell and corresponding quantification of FRET intensity across the cell . Lower panel show the normalised FRET intencity at the furrow (scramble: 76,3 ± 27,6; n=18, Abba-shRNA3: 44,8 ± 17,2; n=15, Abba-shRNA + ABBA-FL: 102.,5 ± 49,7 n=23, Abba-FL: 81,6 ± 38,7,0; n=22; p<0.0001, Abba-R671W: 135,7 ± 62,4 n=15). Error bars represent mean ± s.d. **P < 0.002, ***P < 0.001. Scale bar 25 μm (**A**).

### Identification of *ABBA* missense variant R671W in an individual with neurodevelopmental syndrome

Mutations in genes affecting mitotic progression of neuronal progenitors have been linked to neurodevelopmental disorders^41^. Importantly, we identified a patient carrying a novel heterozygous missense variant in *ABBA* coding *MTSS1l/MTSS2* gene. This mutation (2011C>T(R671W)) affects residues that are conserved from Zebrafish to Human and reside in exon 15 (Figure 6A and B). Consistent with a d*e novo* origin of the variant, it was not found in the parents of the affected individual or in normal controls. This patient presents some characteristic craniofacial dysmorphism as mild deep-set eyes, thickened helices, synophrys, narrow forehead, bitemporal narrowing, small bilateral epicanthal folds, upslanting palpebral fissures, medial flaring of eyebrows, flat midface, V-shaped palate, and dysmorphic ears. Brain magnetic resonance imaging sequences showed the presence of a microcephalic brain associated with a smaller head linked to mild IQ. The patient also suffered from Attention-Deficit/Hyperactivity Disorder (ADHD) (Figure 6C, Supplemental Table 1). Moreover, recent publications found six additional patients carrying the same *ABBA* variant that shared similar brain malformation^42,43^. This variant replaces an arginine to tryptophan that has a large aromatic group that could lead to destabilization of a tertiary structure. Indeed, bioinformatic prediction using AlphaFold predicts (pLDDT) with high 84.4 confidence score that R671 is located in an alpha helical fold close to the C-terminal end of the protein^44^. Furthermore, by using Poly-Phen-2 structure and sequence-based algorithm^23^ to predict the impact of R671W substitution on the structure and function of ABBA, we found a high probability that the mutation is damaging to the protein function (Naïve bayes probability score 0.952) (Figure 6D).

**Figure 6.**
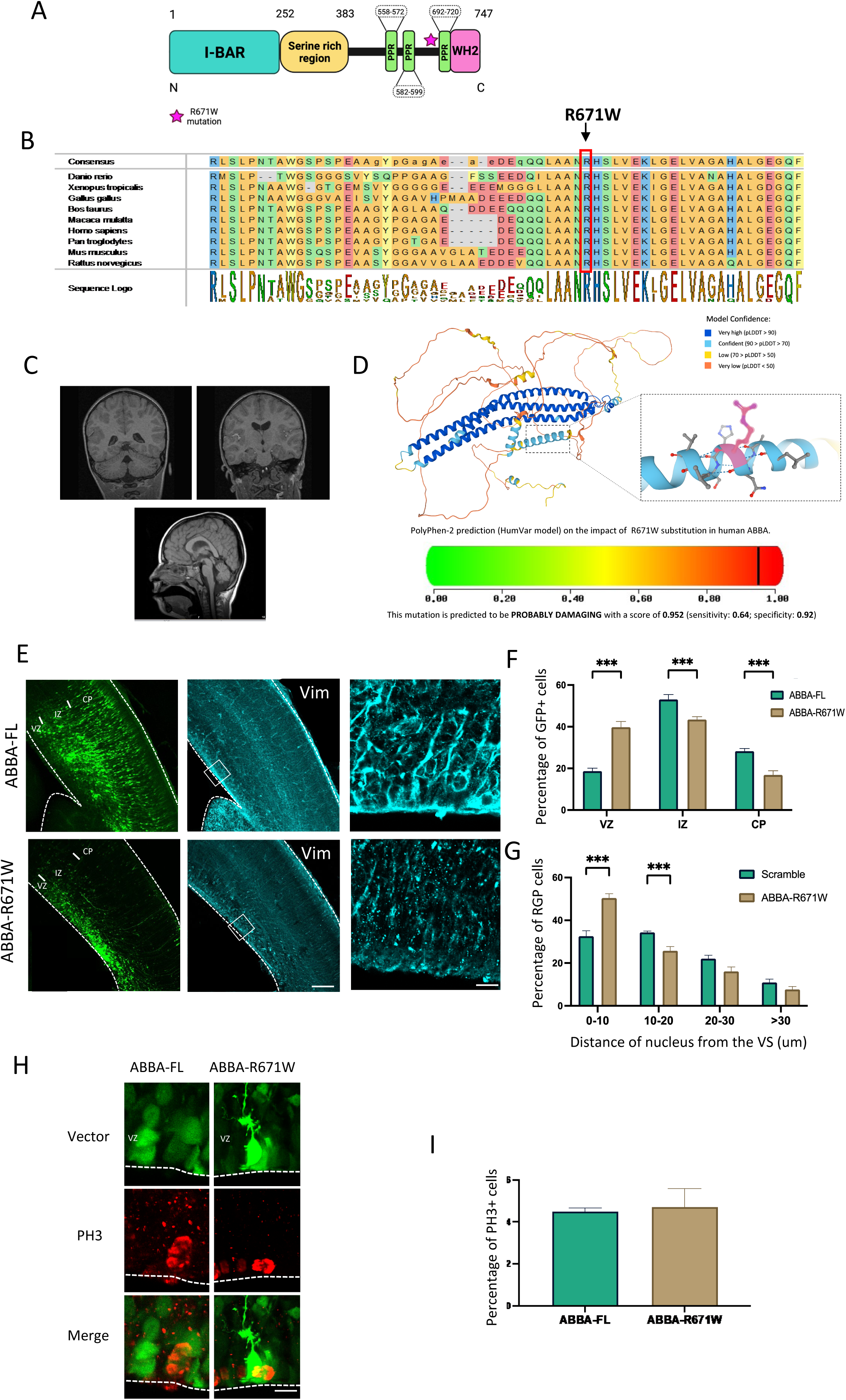
Effect of ABBA R671W human mutation on neuronal migration and mitosis. **A.** ABBA is composed of an N-terminal BAR domain, a serine-rich region, three proline-rich motifs and a C- terminal WH2 domain. **B.** Evolutionary conservation analysis revealed that the Arg671 site is conserved from zebrafish to humans. **C.** Representative brain imaging features of one patient carrying ABBA R671W variant. **D.** 3D structure of ABBA denoting the position of Arginine 571. Lower panel show the high probability of disruption of the α-helix conformation by an Arginine to Tryptophane mutation. **E.** Coronal sections of E17 mice brains electroporated at E14 with cDNAs encoding the human wild-type, ABBA-FL, or the human mutant form of ABBA, ABBA-R671W. Expression of ABBA-R671W results in a defect in neuronal migration as indicated by the accumulation of neurons in the SVZ/lower IZ as well as a disorganization of the radial glial cell fibers using vimentin staining. **F.** Quantification of cell distribution in the cortex at E17 (VZ/SVZ: ABBA-FL 18.67±1.44%, ABBA-R671W 39.77±2.73%; IZ: ABBA-FL 53.07±2.37%, ABBA-R671W 43.40±1.40%; CP: ABBA-FL 28.25±1.25%, ABBA-R671W 16.8±2.03%; n=6) showing a significant increase of mutant cells in SVZ/lower IZ. **G.** Quantification of the distance of RGP nuclei from the ventricular surface (VS) at E17 (0–10: ABBA-FL 32.56±2.58%, ABBA-R671W 50,04±2.04%; 10–20: ABBA-FL 34.37±0,59%, ABBA-R671W 25.76±1.97%; 20–30: ABBA-FL 22.08±1.59%, ABBA-R671W 16.12±2.12%; >30: ABBA-FL 10.99±1.52%, ABBA-R671W 7.71±1.38%; n=8 ABBA-FL and n=6 ABBA-R671W), revealing accumulation at this site for the mutant, but not wild-type ABBA. (VZ/SVZ: p=0.000001: IZ: p=0.0014; CP: p=0.0003; 0–10: p=0.0002; 10–20: p=0.0005; 20–30: p=0.040; >30: p=0.150); mean ± SD. **H.** Brains were fixed at E17 and stained with the mitotic marker Phopho-Histone3 (PH3). **I.** The percentage of PH3-positive nuclei located at the ventricular surface (VS) did not differ in RGP cells expressing ABBA-R671W (ABBA-FL, 4.50±0.17%, n=9; ABBA-R671W, 4.71±0.88%, n=5, P=0.761;). Error bars represent mean ± s.d. **P < 0.01, ***P < 0.001. VZ: ventricular Zone, IZ: Intermediate Zone, CP: Cortical Plate. Scale bars 100μm, 50μm (E), 20μm (H).

### Expression of R671W Human variant results in defects in mitosis and neuronal migration

To test the effects of the human missense variant in mouse brain development, we introduced the human *ABBA* cDNA encoding either wild type or patient mutation R671W into developing mouse brains by *in utero* electroporation at E14 followed by imaging and quantification of targeted cells at E17. Interestingly, whereas wild type ABBA (ABBA-FL) expression showed no defects in radial glial morphology or neuronal migration, the R671W mutant expression resulted in disorganized radial glial fibers and accumulation of multipolar neurons in the SVZ/lower IZ with very few bipolar neurons (Figure 6E). When analyzing the distribution of electroporated neuron progenitors, we observed clear accumulation in the VZ/SVZ (Figure 6F; ABBA-FL 18.67±1.44%; n=8, ABBA-R671W 39.77±2.73% n=6; P=0.000. Supplementary Figure 3B-C). We also found a significant increase in RGP nuclei at the VS (Figure 6G, C ABBA-FL 32.56±2.58%; n=8, ABBA-R671W 50,04±2.04%; n =6; p=0.000001. Supplementary Figure 3B-C) and changes in the distance of the nucleus from the VZ. Interestingly, no difference in accumulation of PH3+ cells was found (Supplemental Figure 4 B and C; Abba-human-WT 4.50 ± 0.17%; n=9, Abba-human-mutation 4.71 ± 1.98%; p=0.76; n=5), indicative of a dysfunction in neuronal maturation but a less prominent impact on cell division. In line with these results the effect of ABBA-R671W on RhoA activation was not significantly different to scramble expressing cells (142 ± 17,9 n=15). The data presented indicate that the abnormal expression of the R671W mutation, rather than the normal protein, leads to abnormal development of the developing cortex. These findings offer evidence suggesting a potential connection between the missense mutation in ABBA discovered in the patient and abnormal brain development. However, it is unlikely that the mechanism behind this phenotype is a result of ABBA-R671W acting in a dominant-negative manner on cell cycle progression.

## Discussion

Abba was initially discovered as a novel regulator of actin and plasma membrane dynamics, specifically in radial glial C6 cells. However, its role in brain radial glial progenitor (RGP) cells in live organisms has remained unclear. In this study, we observed that decreased expression of Abba in radial glia within the cortical ventricular zone leads to impaired radial migration and disrupted organization of radial glia. Notably, knock-down of Abba specifically hindered cytokinesis in RGP cells, resulting in abnormal cell morphology, mitosis, and reduced numbers of glial and neuronal cells. The underlying mechanism behind this effect appears to involve impaired signaling through the actin regulatory protein RhoA, likely through its interaction with Nedd9 during the early and late stages of telophase. In accordance with these findings, we identified a missense variant in a patient with microcephaly and intellectual disability exhibiting similar clinical characteristics. Furthermore, overexpression of the mutant protein in cortical progenitors of mice resulted in phenotypes resembling, although not identical to, the down-regulation of *Abba* in the cortex. The results presented in this study highlight the critical importance of *Abba* in cortical development and contribute to elucidating the potential mechanism underlying abnormal brain development associated with human variants of *ABBA*.

### Radial glial exiting from cytokinesis requires Abba

BAR proteins can interact with PI(4,5)P_2_ -rich membranes through their BAR domain, and several studies indicate that PI(4,5)P_2_ play a central role in cytokinesis. Indeed, it has been reported that in mammalian cells PI(4,5)P_2_ accumulates at the cleavage furrow and recruits membrane proteins required for stability of the furrow because of a role in adhesion between the contractile ring and the plasma membrane. Moreover, overexpression of proteins that bind to PI(4,5)P_2_ perturbs cytokinesis completion, by interfering with adhesion of the plasma membrane to the contractile ring at the furrow, but not ingression of the cleavage furrow^45,46^. Immunocytochemistry of Abba shows a strong accumulation at the cleavage furrow. In addition, in absence of Abba, we observed an accumulation of cells blocked in cytokinesis (Figure 3). these observations and previous findings showing that Abba can interact with PI(4,5)P_2_ -rich membranes through its BAR domain as well as with the actin cytoskeleton through Wasp Homology-2 domain (WH2), lead us suspect that Abba could link the actin contractile ring to the plasma membrane and participate in the mechanism for the invagination of the nuclear membrane during cytokinesis.

### Abba interacts with Nedd9 to regulate RhoA activity

Here we have found that Abba interacts directly with Nedd9 and down regulation of Abba leads to significant changes in RhoA activation. Nedd9 is a focal adhesion protein involved in mitotic entry and cleavage furrow ingression^47^. Similar to the results found in this study, previous results have shown that cells with abnormal expression of Nedd9 remain in G1 subsequently leading to cell apoptosis^37^. This is consistent with the view that Nedd9 triggers cells to enter mitosis. Nedd9 is known to be a positive regulator of RhoA signaling in mitosis. Indeed, activation of RhoA is required for cell rounding at early stages of mitosis and cleavage furrow ingression, and deactivation of RhoA is essential for cell abscission^21^. Moreover, it has been shown that Nedd9 overexpression blocks cells at abscission by maintaining levels of RhoA activation, and decreased Nedd9 expression inhibits cleavage furrow ingression by non-activated RhoA pathway. In light of these results, we hypothesized that during cytokinesis at the cleavage furrow the membrane composition change and is enriched in PI(4,5)P_2_ leading to Abba recruitment and its interaction with Nedd9 which is essential to activate the RhoA pathway cleavage furrow ingression.

### Role of ABBA in neuronal migration and microcephaly

The proper formation of the cerebral cortex relies on the sequential generation of cortical layers, which is highly dependent on the normal production, survival, and migration of neuronal progenitors. Abnormalities in these processes are associated with pathological conditions such as microcephaly, leading to impaired cognitive development. Interestingly, both downregulation of *Abba* and overexpression of the *ABBA* point mutation construct result in a similar accumulation of neuronal progenitors in the ventricular zone (VZ) and subventricular zone (SVZ). This may be attributed to abnormal proliferation of neuronal progenitors caused by disrupted interaction with critical proteins involved in cell cycle progression. This disruption aligns with the hypothesized impact on Nedd9 interaction and reduced RhoA activity observed in conditions of low Abba expression.

One possible explanation for these results could be the disruption of the predicted local tertiary structure. In this scenario, heterozygous expression of the human R671W variant would exert a dominant negative effect on *ABBA*’s role in brain development, leading to microcephaly and cognitive delay. This notion is supported by recent work disclosing additional patient carrying the R671W variant^42^. In the same study the significant neurological phenotypes were observed in a drosophila model where the ortholog of human MTSS2 and MTSS1, mim, was deleted. However, from a clinical genetics’ standpoint, it is unlikely to find patients with the recurrent R671W mutation without any homozygous or compound heterozygous loss-of-function mutations elsewhere in the *ABBA* gene. This could also suggest a gain-of-function effect of the R671W mutation. Supporting this notion, overexpressing ABBA-R671W in cells expressing the wild-type *Abba in this study* did not result in a dominant-negative decrease in RhoA activation, nor did it affect the expression of PH3 *in vivo*. These findings make it plausible to suggest that a mechanism responsible for the phenotype associated with overexpression of the human variant may primarily involve post-cell division processes, such as cell migration.

Although previous research has indicated that Abba is not expressed in mature neurons^20^, more recent studies have shown transient expression of Abba in neurons during conditions of increased neuronal plasticity^13^. While our study primarily focused on Abba’s role in radial glial cell proliferation, the data also indicate an additional role in the migration of immature neurons. This is supported by the observed accumulation of Abba-shRNA3 electroporated cells in the ventricular zone. Additionally, we observed a significant decrease in process length in migrating cortical neurons with downregulated Abba and expressing ABBA-R671W mutant but interestingly dendritic processes were increased in length in neurons overexpressing *ABBA* (Supplemental Figure 5). Given that disorganization of radial glial progenitor cells does not lead to neuronal cell death, these findings suggest that Abba may be involved in additional molecular mechanisms in differentiated neurons. Further experiments are needed to determine whether this phenotype is intrinsic to Abba’s role in migrating neurons or if it is influenced by the disruption of guiding radial glia.

In conclusion, this study provides compelling evidence for the significant role of *Abba* in brain development. We demonstrate that Abba’s involvement in the cell division of radial glial progenitors is a key component of this mechanism. Furthermore, these findings have translational relevance as they contribute to a better understanding of the putative mechanisms underlying human variants associated with microcephaly and developmental disabilities.

**Table 1.** Summary of clinical and imaging phenotypes associated with mutations in ABBA.

**Table 2.** Y2H screening results.

## Data Availability

All data produced in the present study are available upon reasonable request to the authors

## Acknowledgements

We thank Drs Lauren Briere and Gabrielle Lemire for discussion about the data. We thank Biomedicum Imaging Unit (Helsinki, Finland) for their technical assistance with the time-lapse imaging of organotypic cultures. This project was supported by Eranet Neuron III program project ACROBAT, ANR project GABGANG and Academy of Finland grant 341361 to C.R., and by Fondation pour la Recherche Medicale “aide au retour en France” to A.C. Academy of Finland (317038, 319907) and Sigrid Juselius foundation to J.S. and Finnish Cultural Foundation to E.E.

## Author Contributions

A.C conceived the project. A.C. designed and performed most of the experiments, analyzed and interpreted all the data, and prepared figures. A.C. and C.R. wrote the original draft of the manuscript. J.S. designed experiments, interpreted data, edited the manuscript. E.E: performed and analyzed experiments and edited the manuscript. H.K. performed organotypic slices culture and live imaging experiments. All authors read and approved the final manuscript.

## Figures Legends

**Supplemental figure 1. Distribution and directionality of RGP. A.** representative picture of the distribution of RFP expressing cells at E17 and corresponding cortical layer markers Cux1 and Ctip2. **B.** Quantification of dispersion of Vimentin staining similar as in Figure 1 in different conditions (Scramble ; 14.27.4±6 % and Abba- shRNA3 40.8 ±19, n=20 P=0.0078).

**Supplemental figure 2 . Alteration of Abba expression by RNAi**

**A.** Schematic diagram showing the position of small-hairpin RNAs targeting the coding sequence (CDShp) and the 3′UTR of Abba mRNA. **B.** Knockdown of endogenous Abba mRNA expression in rat C6 glioma cells was measured by qPCR 72 h after transfection with CDShp or 3UTRhp. We observed a reduction of Abba mRNA expression by 70% with Abba-shRNA3 compared with corresponding ineffective shRNAs (Abba-shRNA1 and Abba-shRNA2) or the control scramble. Rpl13a was used for normalization. **C.** Western blot analysis revealed that Abba protein levels, 72 h after transfection, were much lower in cells transfected with Abba-shRNA3 compared with those transfected with scramble and with non-transfected (control) cells. α-Tubulin was used for normalization. **D.** We observed a reduction of Abba expression in Rat C6 glioma cells, 72h after transfection with Abba-shRNA3 compared with cells transfected with scramble. E. Interaction between Nedd9 and Abba assessed by Immunoprecipitation from C6 cell homogenates. Error bars represent mean ± s.d. **P < 0.02, ***P < 0.001. Scale bars: 20 µm (D).

**Supplemental figure 3 . Flow cytometry analysis of cells C6 cells.**

**A.** Cells expressing Abba-shRNA3 show accumulation in S-phase and recovery after additional expression of Abba-FL (**G1**: scramble 45,2 ± 1,4 %, Abba-shRNA3 37,4 ± 1,0 %, Abba-shRNA3+Abba-FL 42,8 ± 1.3 %; **S**: scramble 34,3 ± 2,8 %, Abba-shRNA3 48,2 ± 2.7 %, Abba-shRNA3+Abba-FL 38,9 ± 3.4; **G2**: scramble 17,1 ± 2,1 %, Abba-shRNA3 14,4 ± 2,7%, Abba-shRNA3+Abba-FL 15,9 ± 1,9 %; n = 10). Error bars represent mean ± s.d. **P < 0.002, ***P < 0.001. **B.** Quantification of the distance of RGPC nuclei from the ventricular surface (VS) at E17 (**0–10**: scramble 30.19±3.2%, Abba-shRNA3 40.61±2.7%, ABBA-FL 37.81±1.6%, ABBA-R671W 50.40 ±4.9%, Nedd9-shRNA 46.13 ±2.4%; **10–20**: scramble 34.34±1.8%, Abba-shRNA3 21.55±2.4%, ABBA-FL 31.19±2.3%, ABBA-R671W 25.76 ±4.8%, Nedd9-shRNA 28.58±5.8%; **20–30**: scramble 23.41±2.6%, Abba-shRNA3 16.34±2.3%, ABBA-FL 20.40±3.2%, ABBA-R671W 16.11±5.1%, Nedd9-shRNA 18.39±4.8%; **>30**: scramble 12.03±3.3%, Abba-shRNA3 21.49±2.1%, ABBA-FL 10.58±3.2%, ABBA-R671W 7.70±3.4%, Nedd9- shRNA 6.88±2.7%; n=7 scramble and Abba-shRNA3, n=6 Nedd9-shRNA and ABBA-R671W, n=5 ABBA-FL). **C.** quantification of the number of RGPC in the ventricular zone (VZ) at E17 (scramble 126.5±37.8, Abba-shRNA3 48.28±20.9, ABBA-FL 58±25.7, ABBA-R671W 35.16 ±16.7, Nedd9-shRNA 82.66 ±18.4).

**Supplemental figure 4 . Nedd9 shRNA efficiency test in C6 cells and effect on PH3 positive cell *in vivo*.**

**A.** Quantification of Nedd9 mRNA expression in C6 cell after 72h transfection with four different shRNA constructs. Sh3 shows most efficiently decreased Nedd9 expression. **B.** Scramble and Nedd9-shRNA electroporated brains were stained with the late G2/M phase marker Phospho-Histone3 (PH3). **C.** The percentage of PH3-positive nuclei located at the ventricular surface (VS) did not increase in RGP cells expressing Nedd9-shRNA (scramble: 4.50±0.18%, n=9; Nedd9-shRNA: 5.07±0.35%, n=6, P=0,1135;). **D.** C6 cell homogenates were subjected to a direct pulldown assay. Nedd9 co-immunoprecipitated with endogenous Abba indicating the specificity of the interaction. Error bars represent mean ± s.d. **P < 0.002, ***P < 0.001. VZ: ventricular Zone, IZ: Intermediate Zone, CP: Cortical Plate. Scale bars 100μm (A), 20μm (D).

**Supplemental figure 5 . Impact of suppression and overexpression of ABBA on migrating neurons**.

The figure illustrates the variation in the length and total number of processes in cortical neurons electroporated either with Abba-ShRNA3 or ABBA-FL and ABBA mutant (upper panel). Lower panel: Staining with Abba antibody. IZ: Intermediate Zone, CP: Cortical Plate. Scale bars 20μm.

**Supplementary Figure 6. Quantification of Anillin expression changes during 15h live imaging recordings.** A. Shows a representative image Z-stack projection of a single time point. B. Graph showing the results from tracking of number of Anillin expressing cell at each time point during 15h recording was quantified under different conditions (Scramble ; 100.2 ±15 % and Abba-shRNA3 152.6 22, n=20 P=0.036)

**Movie 1: symmetric division**

Related to Figure 3C, top panel. E14 mouse embryonic brain was electroporated with vectors expressing Abba shRNA and RFP. Brain was sectioned at E17 and imaged every 15 minutes. Nucleus of RGP cell apically migrated to the ventricular surface and underwent symmetric division.

**Movie 2: absence of division in Abba depleted progenitor cells**

Related to Figure 3C, bottom panel. E14 mouse embryonic brain was electroporated with vectors expressing Abba shRNA and RFP. Brain was sectioned at E17 and imaged every 15 minutes. Nucleus of RGP cell apically migrated to the apical surface and remains blocked at the ventricular surface for hours without being able to enter mitosis.

