## Supplementary material for "Identification of novel microcephaly-linked protein ABBA that mediates cortical progenitor cell division and corticogenesis through NEDD9-RhoA": table 2

HYBRIGENICS

services

*Results Summary*

### Mouse ABBA-1
### vs Mouse Embryo Brain RP2

#### *Screen Parameters*

| | |
|---|---|
| Nature | cDNA |
| Reference Bait Fragment | Mouse ABBA-1 (1-715) ; hgx2114v1 |
| Prey Library | Mouse Embryo Brain RP2 |
| Vector(s) | pB27 (N-LexA-bait-C fusion) + pB66 (N-GAL4-bait-C fusion) |
| Processed Clones | 15 (pB27) + 269 (pB66) + 87 (pB66) |
| Analyzed Interactions | 91 millions (pB27) + 23.2 millions (pB66) + 8.12 millions (pB66) |
| 3AT Concentration | 0.0 mM (pB27) + 0.0 mM (pB66) + 0.0 mM (pB66) |

#### *Global PBS®*

| | Global PBS (for Interactions represented in the Screen) | Nb | % |
|---|---|---|---|
| A | Very high confidence in the interaction | 0 | 0.0% |
| B | High confidence in the interaction | 1 | 1.0% |
| C | Good confidence in the interaction | 3 | 2.9% |
| D | Moderate confidence in the interaction<br>This category is the most difficult to interpret because it mixes two classes of interactions :<br>- False-positive interactions<br>- Interactions hardly detectable by the Y2H technique (low representation of the mRNA in the library, prey folding, prey toxicity in yeast) | 95 | 93.1% |
| E | Interactions involving highly connected prey domains, warning of non-specific interaction. The threshold for high connectivity is 10 for screens with Human, Mouse, Drosophila and Arabidopsis and 6 for all other organisms. They can be classified in different categories:<br>- Prey proteins that are known to be highly connected due to their biological function<br>- Proteins with a prey interacting domain that contains a known protein interaction motif or a biochemically promiscuous motif | 1 | 1.0% |
| F | Experimentally proven technical artifacts | 2 | 2.0% |
| | Non Appliable | | |
| N/A | The PBS is a score that is automatically computed through algorithms and cannot be attributed for the following reasons :<br>- All the fragments of the same reference CDS are antisens<br>- The 5p sequence is missing<br>- All the fragments of the same reference CDS are either all OOF1 or all OOF2<br>- All the fragments of the same reference CDS lie in the 5' or 3' UTR | | |

#### *Prey Fragment Analysis*

| Symbols | Means |
|---|---|
| | The fragment contains the full length CDS |
| | Fragment is fully in 5' UTR |
| | Fragment is fully in 3' UTR |
| | Fragment contains at least one In Frame STOP codon |
| [NR] | Fragment was found to be non relevant (poor quality, high N density) |
| IF<br>OOF1<br>OOF2 | With regard to the theoretical frame of each corresponding CDS (GeneBank),fragments are cloned in frame (IF) if they are in the same frame as Gal4AD.In general, polypeptides synthesized from OOF fragments are not considered of biological interest, unless found together with another frame. However, some of the proteins expressed from an OOF fragment can be translated in the correct frame,due to the existence of natural frame-shift events during translation in yeast |
| ?? | Unidentified frame when :<br>- The clone sequence is antisense<br>- The 5p sequence is missing |
| N | Antisens |
| Start...Stop | Position of the 5p and 3p prey fragment ends, relative to the position of the ATG start codon (A=0) |

| Clone Name | Type Seq | Gene Name (Best Match) | Start..Stop (nt) | Frame | Sens | %Id 5p | %Id 3p | PBS |
|---|---|---|---|---|---|---|---|---|
| pB66_A-88 | 5p/3p | Mus musculus - ABBA-1 | -52..780 | IF | | 99.0 | 99.8 | D |
| pB66_A-185 | 5p/3p | Mus musculus - AU016693 | 3053..3797 | OOF2 | | 99.8 | 99.8 | N/A |
| pB66_A-261 | 3p | Mus musculus - Atf2 | ..2101 | ?? | N | | ALU 98.1 | N/A |
| pB66_B-93 | 5p/3p | Mus musculus - BC018245 | 2873..1560 | ?? | N | 69.1 | 99.1 | N/A |
| pB66_A-76 | 5p/3p | Mus musculus - BC018245 | 2873..1560 | ?? | N | 78.4 | 100.0 | N/A |
| pB66_A-165 | 5p/3p | Mus musculus - BC066049 | 2560..2089 | ?? | N | 100.0 | 100.0 | N/A |
| pB66_A-120 | 5p/3p | Mus musculus - BC066049 | 2569..2258 | ?? | N | 99.7 | 99.7 | N/A |
| pB66_A-18 | 5p/3p | Mus musculus - BC066049 | 2569..2258 | ?? | N | 99.7 | 99.7 | N/A |
| pB66_A-169 | 5p/3p | Mus musculus - BC086774 | 1438..2127 | OOF1 | | 85.2 | 99.3 | N/A |
| pB66_A-254 | 3p | Mus musculus - Cdca5 | ..49 | ?? | N | | 78.5 | N/A |
| pB66_B-80 | 5p/3p | Mus musculus - Cdca5 | 465..-48 | ?? | N | 92.7 | 90.9 | N/A |
| pB66_A-212 | 5p/3p | Mus musculus - Cdca5 | 465..-101 | ?? | N | 93.7 | 99.6 | N/A |
| pB66_A-202 | 5p/3p | Mus musculus - Cdca5 | 550..30 | ?? | N | 73.6 | 100.0 | N/A |
| pB66_B-24 | 5p/3p | Mus musculus - Cdca5 | 551..49 | ?? | N | 92.0 | 100.0 | N/A |
| pB66_A-209 | 5p/3p | Mus musculus - Cdca5 | 552..30 | ?? | N | 91.4 | 99.8 | N/A |
| pB66_A-174 | 5p/3p | Mus musculus - Cdca5 | 552..30 | ?? | N | 100.0 | 100.0 | N/A |
| pB66_A-173 | 5p/3p | Mus musculus - Cdca5 | 552..30 | ?? | N | 100.0 | 100.0 | N/A |
| pB66_A-176 | 5p/3p | Mus musculus - Cdca5 | 552..30 | ?? | N | 100.0 | 100.0 | N/A |
| pB66_B-53 | 5p/3p | Mus musculus - Cdca5 | 552..30 | ?? | N | 99.2 | 100.0 | N/A |
| pB66_A-143 | 5p/3p | Mus musculus - Cdca5 | 552..30 | ?? | N | 100.0 | 100.0 | N/A |
| pB66_A-205 | 5p/3p | Mus musculus - Cdca5 | 552..30 | ?? | N | 100.0 | 100.0 | N/A |
| pB66_B-89 | 5p/3p | Mus musculus - Cdca5 | 552..30 | ?? | N | 100.0 | 100.0 | N/A |
| pB66_A-225 | 5p/3p | Mus musculus - Cdca5 | 552..30 | ?? | N | 100.0 | 100.0 | N/A |
| pB66_A-177 | 5p/3p | Mus musculus - Cdca5 | 552..30 | ?? | N | 100.0 | 100.0 | N/A |
| pB66_A-231 | 5p/3p | Mus musculus - Cdca5 | 552..49 | ?? | N | 99.6 | 100.0 | N/A |
| pB66_B-30 | 5p/3p | Mus musculus - Cdca5 | 552..49 | ?? | N | 99.6 | 99.8 | N/A |
| pB66_B-41 | 5p/3p | Mus musculus - Cdca5 | 552..49 | ?? | N | 99.2 | 100.0 | N/A |
| pB66_A-223 | 5p/3p | Mus musculus - Cdca5 | 552..49 | ?? | N | 98.1 | 100.0 | N/A |
| pB66_A-115 | 5p/3p | Mus musculus - Cdca5 | 552..49 | ?? | N | 100.0 | 100.0 | N/A |

| Clone Name | Type Seq | Gene Name (Best Match) | Start..Stop (nt) | Frame | Sens | %Id 5p | %Id 3p | PBS |
|---|---|---|---|---|---|---|---|---|
| pB66_A-70 | 5p/3p | Mus musculus - Cdca5 | 552..49 | ?? | N | 100.0 | 100.0 | N/A |
| pB66_A-69 | 5p/3p | Mus musculus - Cdca5 | 552..49 | ?? | N | 100.0 | 100.0 | N/A |
| pB66_A-51 | 5p/3p | Mus musculus - Cdca5 | 552..49 | ?? | N | 100.0 | 100.0 | N/A |
| pB66_A-30 | 5p/3p | Mus musculus - Cdca5 | 552..49 | ?? | N | 100.0 | 100.0 | N/A |
| pB66_A-196 | 5p/3p | Mus musculus - Cdca5 | 552..49 | ?? | N | 93.2 | 99.6 | N/A |
| pB66_A-127 | 5p/3p | Mus musculus - Cdca5 | 552..30 | ?? | N | 100.0 | 100.0 | N/A |
| pB66_A-162 | 5p/3p | Mus musculus - Cdca5 | 555..-33 | ?? | N | 100.0 | 100.0 | N/A |
| pB66_A-179 | 5p/3p | Mus musculus - Cdca5 | 558..74 | ?? | N | 100.0 | 100.0 | N/A |
| pB66_A-106 | 5p/3p | Mus musculus - Cdca5 | 558..-23 | ?? | N | 100.0 | 100.0 | N/A |
| pB66_A-262 | 5p/3p | Mus musculus - Cdca5 | 558..-23 | ?? | N | 99.1 | 100.0 | N/A |
| pB66_A-35 | 5p/3p | Mus musculus - Cdca5 | 561..-29 | ?? | N | 100.0 | 100.0 | N/A |
| pB66_A-259 | 5p/3p | Mus musculus - Cdca5 | 561..-29 | ?? | N | 89.5 | 99.7 | N/A |
| pB66_A-112 | 5p/3p | Mus musculus - Cdca5 | 561..-29 | ?? | N | 100.0 | 100.0 | N/A |
| pB66_A-249 | 5p/3p | Mus musculus - Cdca5 | 657..325 | ?? | N | 99.4 | 100.0 | N/A |
| pB66_A-110 | 5p/3p | Mus musculus - Cdca5 | 657..325 | ?? | N | 100.0 | 100.0 | N/A |
| pB66_B-26 | 3p | Mus musculus - Cdca5 | 1057..554 | ?? | N | | 100.0 | N/A |
| pB66_A-215 | 3p | Mus musculus - Cdca5 | 1057..554 | ?? | N | | 100.0 | N/A |
| pB66_A-217 | 3p | Mus musculus - Cdca5 | 1057..554 | ?? | N | | 100.0 | N/A |
| pB66_A-226 | 3p | Mus musculus - Cdca5 | 1068..554 | ?? | N | | 100.0 | N/A |
| pB66_A-198 | 3p | Mus musculus - Cdca5 | 1076..554 | ?? | N | | 100.0 | N/A |
| pB66_A-272 | 3p | Mus musculus - Cdca5 | 1078..554 | ?? | N | | 99.8 | N/A |
| pB66_A-283 | 3p | Mus musculus - Cdca5 | 1140..560 | ?? | N | | 99.5 | N/A |
| pB66_A-166 | 3p | Mus musculus - Cep70 | ..1335 | ?? | N | | 98.8 | N/A |
| pB66_B-91 | 5p/3p | Mus musculus - Cep70 | 2245..1335 | ?? | N | 99.0 | 98.6 | N/A |
| pB66_B-33 | 5p/3p | Mus musculus - Cep70 | 2245..1335 | ?? | N | 98.1 | 98.8 | N/A |
| pB66_B-76 | 5p/3p | Mus musculus - Cep70 | 2245..1335 | ?? | N | 99.0 | 98.6 | N/A |
| pB66_A-83 | 5p/3p | Mus musculus - Cep70 | 2245..1335 | ?? | N | 99.1 | 99.5 | N/A |
| pB66_A-94 | 5p/3p | Mus musculus - Cep70 | 2245..1335 | ?? | N | 99.2 | 99.3 | N/A |
| pB66_A-164 | 5p/3p | Mus musculus - Cep70 | 2245..1335 | ?? | N | 99.2 | 99.0 | N/A |
| pB66_A-218 | 5p/3p | Mus musculus - Cep70 | 2245..1335 | ?? | N | 97.9 | 98.5 | N/A |
| pB66_A-27 | 5p/3p | Mus musculus - Cep70 | 2245..1335 | ?? | N | 99.4 | 99.0 | N/A |
| pB66_A-159 | 5p/3p | Mus musculus - Cep70 | 2245..1335 | ?? | N | 98.8 | 93.2 | N/A |
| pB66_B-72 | 5p/3p | Mus musculus - Cep70 | 2245..1335 | ?? | N | 98.4 | 97.5 | N/A |
| pB66_B-38 | 5p/3p | Mus musculus - Cep70 | 2218..1160 | ?? | N | 97.4 | 81.6 | N/A |
| pB66_B-16 | 5p/3p | Mus musculus - Cops5 | 78..827 | IF | | 74.5 | 100.0 | F |
| pB66_B-55 | 5p/3p | Mus musculus - Csnk1g3 | 1598..2110 | OOF2 | | 100.0 | 100.0 | N/A |
| pB66_A-211 | 5p/3p | Mus musculus - D15Wsu75e | 410..1300 | OOF2 | | 97.9 | 99.5 | N/A |
| pB66_B-46 | 5p/3p | Mus musculus - D15Wsu75e | 452..1000 | OOF2 | | 100.0 | 100.0 | N/A |
| pB66_B-52 | 5p/3p | Mus musculus - D15Wsu75e | 473..1040 | OOF2 | | 99.8 | 100.0 | N/A |
| pB66_B-54 | 5p/3p | Mus musculus - D15Wsu75e | 473..853 | OOF2 | | 99.0 | 100.0 | N/A |
| pB66_A-154 | 5p/3p | Mus musculus - D15Wsu75e | 473..853 | OOF2 | | 100.0 | 100.0 | N/A |
| pB66_B-77 | 5p/3p | Mus musculus - D15Wsu75e | 473..853 | OOF2 | | 89.8 | 100.0 | N/A |
| pB66_B-44 | 5p/3p | Mus musculus - D15Wsu75e | 473..853 | OOF2 | | 96.1 | 100.0 | N/A |
| pB66_B-90 | 5p/3p | Mus musculus - D15Wsu75e | 479..847 | OOF2 | | 98.6 | 100.0 | N/A |
| pB66_B-82 | 5p/3p | Mus musculus - D15Wsu75e | 479..847 | OOF2 | | 96.5 | 100.0 | N/A |
| pB66_A-22 | 5p/3p | Mus musculus - D15Wsu75e | 479..847 | OOF2 | | 100.0 | 100.0 | N/A |
| pB66_A-40 | 5p/3p | Mus musculus - D15Wsu75e | 479..847 | OOF2 | | 98.9 | 98.1 | N/A |
| pB66_A-11 | 5p/3p | Mus musculus - D15Wsu75e | 479..847 | OOF2 | | 100.0 | 100.0 | N/A |
| pB66_A-265 | 5p/3p | Mus musculus - Dach2 | 2398..1352 | ?? | N | 86.2 | 97.1 | N/A |
| pB66_B-71 | 5p/3p | Mus musculus - Dach2 | 2399..1352 | ?? | N | 98.5 | 95.5 | N/A |
| pB66_B-19 | 5p/3p | Mus musculus - Dach2 | 2399..1352 | ?? | N | 99.0 | 96.1 | N/A |

| Clone Name | Type Seq | Gene Name (Best Match) | Start..Stop (nt) | Frame | Sens | %Id 5p | %Id 3p | PBS |
|---|---|---|---|---|---|---|---|---|
| pB66_B-84 | 5p | Mus musculus - Dach2 | 2399 | ?? | N | 96.4 | | N/A |
| pB66_A-178 | 5p/3p | Mus musculus - Dach2 | 2399..1352 | ?? | N | 99.2 | 95.8 | N/A |
| pB66_A-234 | 5p/3p | Mus musculus - Dach2 | 2399..1352 | ?? | N | 98.8 | 95.6 | N/A |
| pB66_A-28 | 5p/3p | Mus musculus - Dach2 | 2399..1352 | ?? | N | 98.7 | 96.6 | N/A |
| pB66_A-17 | 5p/3p | Mus musculus - Dach2 | 2399..1352 | ?? | N | 98.7 | 97.1 | N/A |
| pB66_A-21 | 5p/3p | Mus musculus - Dgcr8 | 1254..716 | ?? | N | 100.0 | 99.8 | N/A |
| pB66_B-17 | 5p/3p | Mus musculus - Dip2b | 1796..868 | ?? | N | 81.4 | 83.3 | B |
| pB66_A-253 | 3p | Mus musculus - Fbxw11 | ..1828 | ?? | | | 99.7 | B |
| pB27_A-14 | 5p/3p | Mus musculus - Fbxw11 | 699..1844 | IF | | 99.4 | 100.0 | B |
| pB27_A-10 | 5p/3p | Mus musculus - Fbxw11 | 699..1844 | IF | | 99.7 | 100.0 | B |
| pB27_A-9 | 5p/3p | Mus musculus - Fbxw11 | 699..1844 | IF | | 99.3 | 100.0 | B |
| pB27_A-17 | 5p/3p | Mus musculus - Fbxw11 | 699..1844 | IF | | 99.6 | 99.3 | B |
| pB27_A-4 | 5p/3p | Mus musculus - Fbxw11 | 699..1828 | IF | | 99.4 | 99.7 | B |
| pB27_A-16 | 5p/3p | Mus musculus - Fbxw11 | 711..1810 | IF | | 100.0 | 100.0 | B |
| pB27_A-11 | 5p/3p | Mus musculus - Fbxw11 | 711..1810 | IF | | 98.8 | 99.8 | B |
| pB27_A-18 | 5p/3p | Mus musculus - Fbxw11 | 711..1810 | IF | | 99.6 | 99.8 | B |
| pB66_A-229 | 5p/3p | Mus musculus - Fbxw11 | 711..1810 | IF | | 95.5 | 100.0 | B |
| pB27_A-7 | 5p/3p | Mus musculus - Fbxw11 | 711..1810 | IF | | 99.7 | 100.0 | B |
| pB27_A-12 | 5p/3p | Mus musculus - Glrx3 | 363..960 | IF | | 89.3 | 89.9 | D |
| pB66_B-25 | 5p/3p | Mus musculus - HTC_FLI | 16..398 | OOF1 | | 89.3 | 100.0 | N/A |
| pB66_A-213 | 5p/3p | Mus musculus - HTC_FLI | 16..398 | OOF1 | | 99.2 | 100.0 | N/A |
| pB66_B-83 | 5p/3p | Mus musculus - HTC_FLI | 16..398 | OOF1 | | 100.0 | 100.0 | N/A |
| pB66_A-183 | 5p/3p | Mus musculus - HTC_FLI | 16..398 | OOF1 | | 100.0 | 100.0 | N/A |
| pB66_A-180 | 5p/3p | Mus musculus - HTC_FLI | 16..398 | OOF1 | | 100.0 | 100.0 | N/A |
| pB66_A-102 | 5p/3p | Mus musculus - HTC_FLI | 16..398 | OOF1 | | 100.0 | 100.0 | N/A |
| pB66_A-57 | 5p/3p | Mus musculus - HTC_FLI | 16..398 | OOF1 | | 100.0 | 100.0 | N/A |
| pB66_B-45 | 5p/3p | Mus musculus - Isca1 | 1141..1836 | OOF1 | | 99.6 | 99.8 | N/A |
| pB66_B-3 | 5p/3p | Mus musculus - Isca1 | 1141..1836 | OOF1 | | 99.9 | 100.0 | N/A |
| pB66_A-117 | 5p/3p | Mus musculus - Isca1 | 1141..1682 | OOF1 | | 100.0 | 100.0 | N/A |
| pB66_A-9 | 5p/3p | Mus musculus - Isca1 | 1141..1682 | OOF1 | | 100.0 | 100.0 | N/A |
| pB66_A-237 | 5p/3p | Mus musculus - Isca1 | 1141..1682 | OOF1 | | 99.8 | 97.6 | N/A |
| pB66_A-150 | 5p/3p | Mus musculus - Isca1 | 1141..1682 | OOF1 | | 99.3 | 100.0 | N/A |
| pB66_A-258 | 5p/3p | Mus musculus - Isca1 | 1141..1682 | OOF1 | | 99.8 | 93.7 | N/A |
| pB66_A-46 | 5p/3p | Mus musculus - Isca1 | 1141..1836 | OOF1 | | 99.7 | 99.8 | N/A |
| pB66_A-84 | 5p/3p | Mus musculus - Isca1 | 1141..1836 | OOF1 | | 99.8 | 100.0 | N/A |
| pB66_A-232 | 5p | Mus musculus - Isca1 | 1141 | OOF1 | | 97.1 | | N/A |
| pB66_A-247 | 5p/3p | Mus musculus - Isca1 | 1141..1682 | OOF1 | | 82.5 | 99.8 | N/A |
| pB66_A-5 | 5p/3p | Mus musculus - Isca1 | 1141..1836 | OOF1 | | 97.3 | 96.9 | N/A |
| pB66_A-238 | 5p/3p | Mus musculus - Isca1 | 1142..1682 | OOF2 | | 78.0 | 100.0 | N/A |
| pB66_A-149 | 5p/3p | Mus musculus - Isca1 | 1198..1470 | OOF1 | | 98.2 | 100.0 | N/A |
| pB66_A-53 | 5p/3p | Mus musculus - Isca1 | 1198..1470 | OOF1 | | 99.3 | 100.0 | N/A |
| pB66_B-8 | 5p/3p | Mus musculus - Isca1 | 1198..1470 | OOF1 | | 99.3 | 96.0 | N/A |
| pB66_A-107 | 5p/3p | Mus musculus - Isca1 | 1198..1470 | OOF1 | | 100.0 | 100.0 | N/A |
| pB66_A-60 | 5p/3p | Mus musculus - Isca1 | 1198..1470 | OOF1 | | 100.0 | 100.0 | N/A |
| pB66_A-191 | 5p/3p | Mus musculus - Isca1 | 1198..1470 | OOF1 | | 100.0 | 99.6 | N/A |
| pB66_A-90 | 5p/3p | Mus musculus - Kalrn | 105..730 | IF | | 99.8 | 99.5 | D |
| pB66_B-40 | 5p/3p | Mus musculus - LOC100042092 | 1676..1338 | ?? | N | 92.0 | 97.6 | N/A |
| pB66_A-85 | 5p/3p | Mus musculus - LOC100042229 | -1922..-1248 | OOF2 | | 79.5 | 92.5 | N/A |

| Clone Name | Type Seq | Gene Name (Best Match) | Start..Stop (nt) | Frame | Sens | %Id 5p | %Id 3p | PBS |
|---|---|---|---|---|---|---|---|---|
| pB66_A-65 | 5p/3p | Mus musculus - LOC100045522 | 4321..4843 | OOF1 | | 99.8 | 85.7 | N/A |
| pB66_A-138 | 5p/3p | Mus musculus - LOC668039 | 4733..3456 | ?? | N | 98.8 | 99.7 | N/A |
| pB66_A-228 | 5p/3p | Mus musculus - Lrrn1 | 1922..1188 | ?? | N | 93.6 | 99.7 | N/A |
| pB66_A-152 | 5p/3p | Mus musculus - Lrrn1 | 1922..1188 | ?? | N | 100.0 | 99.7 | N/A |
| pB66_A-116 | 5p/3p | Mus musculus - Lrrn1 | 1922..1188 | ?? | N | 100.0 | 100.0 | N/A |
| pB66_A-195 | 5p/3p | Mus musculus - Mll1 | 16440..15614 | ?? | N | 95.0 | 98.0 | N/A |
| pB66_A-182 | 5p/3p | Mus musculus - Mtss1 | -67..777 | IF | | 98.0 | 97.9 | D |
| pB66_B-69 | 5p/3p | Mus musculus - Napg | 1228..1803 | OOF1 | | 99.7 | 100.0 | N/A |
| pB66_A-142 | 5p/3p | Mus musculus - Napg | 1228..1803 | OOF1 | | 99.0 | 100.0 | N/A |
| pB66_A-157 | 5p/3p | Mus musculus - Nedd9 | 2415..3591 | IF | | 99.2 | 99.5 | C |
| pB66_B-22 | 5p/3p | Mus musculus - Nedd9 | 2463..2937 | IF | | 96.8 | 100.0 | C |
| pB66_A-252 | 5p/3p | Mus musculus - Nedd9 | 2463..2937 | IF | | 100.0 | 100.0 | C |
| pB66_A-263 | 5p/3p | Mus musculus - Nedd9 | 2463..2937 | IF | | 92.7 | 95.7 | C |
| pB66_B-62 | 5p/3p | Mus musculus - Ngfr | 1911..1336 | ?? | N | 83.5 | 99.0 | N/A |
| pB66_A-93 | 5p/3p | Mus musculus - Nrxn3 | 3796..4338 | OOF1 | | 99.1 | 100.0 | N/A |
| pB66_A-248 | 5p/3p | Mus musculus - Nrxn3 | 3796..4338 | OOF1 | | 91.4 | 97.8 | N/A |
| pB66_A-13 | 5p/3p | Mus musculus - Nrxn3 | 3796..4338 | OOF1 | | 98.7 | 100.0 | N/A |
| pB66_B-94 | 5p/3p | Mus musculus - Nrxn3 | 3796..4338 | OOF1 | | 95.9 | 99.8 | N/A |
| pB66_B-64 | 5p/3p | Mus musculus - Nrxn3 | 3796..4338 | OOF1 | | 96.3 | 98.3 | N/A |
| pB66_B-34 | 5p/3p | Mus musculus - Nrxn3 | 3796..4338 | OOF1 | | 98.2 | 100.0 | N/A |
| pB66_A-50 | 5p/3p | Mus musculus - Nrxn3 | 3796..4338 | OOF1 | | 95.9 | 100.0 | N/A |
| pB66_A-128 | 5p/3p | Mus musculus - Nrxn3 | 3796..4338 | OOF1 | | 98.5 | 100.0 | N/A |
| pB66_A-34 | 5p/3p | Mus musculus - Nrxn3 | 3796..4338 | OOF1 | | 99.3 | 100.0 | N/A |
| pB66_B-11 | 5p/3p | Mus musculus - Nrxn3 | 3796..4338 | OOF1 | | 99.4 | 99.8 | N/A |
| pB66_A-222 | 5p/3p | Mus musculus - Nsd1 | 4839..5813 | IF | | 98.5 | 99.8 | D |
| pB27_A-6 | 5p/3p | Mus musculus - Nsd1 | 4839..5813 | IF | | 99.9 | 100.0 | D |
| pB66_A-194 | 5p/3p | Mus musculus - Otx2 | 51..283 | IF | | 90.6 | 99.1 | C |
| pB66_A-278 | 5p/3p | Mus musculus - Otx2 | 51..283 | IF | | 95.7 | 97.9 | C |
| pB66_A-250 | 5p/3p | Mus musculus - Otx2 | 51..283 | IF | | 97.0 | 99.6 | C |
| pB66_A-168 | 5p/3p | Mus musculus - Otx2 | 54..283 | IF | | 92.5 | 99.6 | C |
| pB66_A-276 | 5p/3p | Mus musculus - Pax3 | 397..927 | OOF1 | | 82.5 | 93.2 | N/A |
| pB66_A-245 | 3p | Mus musculus - Pax6 | ..1054 | ?? | | | 99.5 | F |
| pB66_B-61 | 3p | Mus musculus - Pax6 | ..933 | ?? | | | 99.8 | F |
| pB66_B-51 | 3p | Mus musculus - Pax6 | ..933 | ?? | | | 99.2 | F |
| pB66_B-47 | 3p | Mus musculus - Pax6 | ..858 | ?? | | | 99.7 | F |
| pB66_A-269 | 3p | Mus musculus - Pax6 | ..858 | ?? | | | 99.1 | F |
| pB66_A-47 | 5p/3p | Mus musculus - Pax6 | 30..890 | IF | | 93.7 | 99.1 | F |
| pB66_B-35 | 5p/3p | Mus musculus - Pax6 | 42..858 | IF | | 93.7 | 98.4 | F |
| pB66_A-134 | 5p/3p | Mus musculus - Pax6 | 105..881 | IF | | 94.0 | 100.0 | F |
| pB66_A-155 | 5p/3p | Mus musculus - Pax6 | 177..1005 | IF | | 99.1 | 97.8 | F |
| pB66_A-75 | 5p/3p | Mus musculus - Pax6 | 225..933 | IF | | 99.6 | 99.5 | F |
| pB66_A-216 | 3p | Mus musculus - Phldb2 | ..3592 | ?? | N | | 98.8 | N/A |
| pB66_A-91 | 5p/3p | Mus musculus - Phldb2 | 4723..3592 | ?? | N | 97.9 | 99.3 | N/A |
| pB66_A-268 | 5p/3p | Mus musculus - Phldb2 | 4723..3592 | ?? | N | 95.1 | 98.9 | N/A |
| pB66_A-24 | 5p/3p | Mus musculus - Ptprs | 5870..6218 | OOF2 | | 99.1 | 99.7 | N/A |
| pB66_A-273 | 3p | Mus musculus - Ranbp9 | ..1351 | ?? | | | 99.8 | N/A |
| pB66_B-10 | 5p/3p | Mus musculus - Rfk | 1494..788 | ?? | N | 86.2 | ALU 94.1 | N/A |
| pB66_A-54 | 5p/3p | Mus musculus - Rsad1 | 2650..3439 | OOF1 | | 99.4 | 99.4 | N/A |

| Clone Name | Type Seq | Gene Name (Best Match) | Start..Stop (nt) | | Frame | Sens | %Id 5p | %Id 3p | PBS |
|---|---|---|---|---|---|---|---|---|---|
| pB66_B-79 | 5p/3p | Mus musculus - Sephs2 | 1764..2070 | ✗ | IF | | ALU 95.7 | 64.1 | N/A |
| pB66_B-39 | 5p/3p | Mus musculus - Sephs2 | 1764..2070 | ✗ | IF | | ALU 95.4 | ALU 65.3 | N/A |
| pB66_A-78 | 5p/3p | Mus musculus - Sephs2 | 1764..2069 | ✗ | IF | | ALU 99.3 | ALU 64.1 | N/A |
| pB66_A-108 | 5p/3p | Mus musculus - Sqol2 | 24..552 | | IF | | 100.0 | 100.0 | D |
| pB66_B-28 | 5p/3p | Mus musculus - Slc2a3 | 2167..2754 | ✗ | OOF1 | | 91.2 | 99.8 | N/A |
| pB66_B-88 | 5p/3p | Mus musculus - Tbl1xr1 | 3025..3490 | ✗ | OOF1 | | 92.5 | 99.6 | N/A |
| pB66_A-203 | 5p/3p | Mus musculus - Tbl1xr1 | 3025..3490 | ✗ | OOF1 | | 92.3 | 100.0 | N/A |
| pB66_A-87 | 5p/3p | Mus musculus - Tbl1xr1 | 3025..3490 | ✗ | OOF1 | | 99.6 | 100.0 | N/A |
| pB66_A-114 | 5p/3p | Mus musculus - Tbl1xr1 | 3025..3490 | ✗ | OOF1 | | 95.0 | 100.0 | N/A |
| pB66_A-32 | 5p/3p | Mus musculus - Tbl1xr1 | 3025..3490 | ✗ | OOF1 | | 100.0 | 100.0 | N/A |
| pB27_A-13 | 5p/3p | Mus musculus - Trim28 | 1021..509 | | ?? | N | 100.0 | 100.0 | N/A |
| pB66_B-75 | 5p | Mus musculus - Uap1 | 390..-18 | | ?? | N | 93.2 | | N/A |
| pB66_B-73 | 3p | Mus musculus - Uap1 | 806..393 | | ?? | N | | 99.5 | N/A |
| pB66_A-210 | 5p/3p | Mus musculus - Whsc1 | 424..-118 | | ?? | N | 97.8 | 97.6 | N/A |
| pB66_A-190 | 5p/3p | Mus musculus - Whsc1 | 424..-118 | | ?? | N | 97.1 | 97.6 | N/A |
| pB66_B-29 | 5p/3p | Mus musculus - Whsc1 | 424..134 | | ?? | N | 91.3 | 98.6 | N/A |
| pB66_A-146 | 5p/3p | Mus musculus - Whsc1 | 424..134 | | ?? | N | 100.0 | 100.0 | N/A |
| pB66_A-96 | 5p/3p | Mus musculus - Whsc1 | 424..-118 | | ?? | N | 97.8 | 97.6 | N/A |
| pB66_A-92 | 5p/3p | Mus musculus - Whsc1 | 424..134 | | ?? | N | 100.0 | 100.0 | N/A |
| pB66_A-97 | 5p/3p | Mus musculus - Whsc1 | 424..-227 | | ?? | N | 87.5 | 86.2 | N/A |
| pB66_A-284 | 5p/3p | Mus musculus - Wwtr1 | 1913..2759 | ✗ | OOF2 | | 92.4 | 98.9 | N/A |
| pB66_A-188 | 5p/3p | Mus musculus - Wwtr1 | 1913..2759 | ✗ | OOF2 | | 94.7 | 98.8 | N/A |
| pB66_A-181 | 5p/3p | Mus musculus - Wwtr1 | 1913..2759 | ✗ | OOF2 | | 88.4 | 99.5 | N/A |
| pB27_A-5 | 5p/3p | Mus musculus - Zbtb16 | 1326..2153 | ✗ | IF | | 98.5 | 99.3 | E |
| pB27_A-8 | 5p/3p | Mus musculus - Zbtb16 | 1326..2153 | ✗ | IF | | 99.4 | 97.9 | E |
| pB27_A-15 | 5p/3p | Mus musculus - Zbtb16 | 1326..2153 | ✗ | IF | | 95.9 | 71.2 | E |
| pB66_A-61 | 5p/3p | Mus musculus - Zmym4 | 1576 | | OOF1 | | 65.5 | 99.5 | N/A |
| pB66_A-282 | 5p/3p | Mus musculus - Zmym4 | 1576 | | OOF1 | | 66.7 | 93.2 | N/A |
| pB66_A-38 | 5p/3p | Mus musculus - Zmym4 | 4456..3078 | | ?? | N | 94.7 | 99.8 | N/A |
| pB66_A-126 | 5p/3p | Mus musculus - Znf512b | 5540..5124 | | ?? | N | 100.0 | 100.0 | N/A |
| pB66_A-255 | 3p | Mus musculus - No Match (no match found in GenBank) | ..560 [NR] | | ?? | | | 100.0 | N/A |
| pB66_B-36 | 5p/3p | Mus musculus - No Match (no match found in GenBank) | | | ?? | | 100.0 | 100.0 | N/A |
| pB66_A-266 | 5p/3p | Mus musculus - GenMatch GID: 47825183 | -1 | | IF | | 80.5 | 94.5 | D |
| pB66_B-95 | 5p/3p | Mus musculus - GenMatch GID: 47825183 | -1 | | IF | | 91.0 | 93.8 | D |
| pB66_B-58 | 5p/3p | Mus musculus - GenMatch GID: 47825183 | -1 | | IF | | 68.1 | 94.1 | D |
| pB66_A-71 | 3p | Mus musculus - GenMatch GID: 56236271 | ..568 | | ?? | | | 100.0 | N/A |
| pB66_A-241 | 5p/3p | Mus musculus - GenMatch GID: 34787446 | -1..497 | ✗ | IF | | 100.0 | 82.2 | D |
| pB66_A-251 | 5p/3p | Mus musculus - GenMatch GID: 26006611 | -1..478 | ✗ | IF | | 100.0 | 75.4 | D |
| pB66_A-279 | 5p/3p | Mus musculus - GenMatch GID: 77404072 | -1..500 | ✗ | IF | | 100.0 | 89.8 | D |
| pB66_A-281 | 5p/3p | Mus musculus - GenMatch GID: 24459826 | -1..393 | ✱ ✗ | IF | | 100.0 | 89.3 | D |
| pB66_B-12 | 5p/3p | Mus musculus - GenMatch GID: 77404072 | -1..502 | ✱ ✗ | IF | | 99.4 | 100.0 | D |

| Clone Name | Type Seq | Gene Name (Best Match) | Start..Stop (nt) | Frame | Sens | %Id 5p | %Id 3p | PBS |
|---|---|---|---|---|---|---|---|---|
| pB66_A-161 | 5p/3p | Mus musculus - GenMatch GID: 77404072 | -1..502 | IF | | 99.8 | 99.8 | D |
| pB66_B-4 | 5p/3p | Mus musculus - GenMatch GID: 33285224 | -1..1006 | IF | | 100.0 | 92.5 | D |
| pB66_A-89 | 5p/3p | Mus musculus - GenMatch GID: 20068727 | -1 | IF | | 100.0 | 82.6 | D |
| pB66_A-26 | 5p/3p | Mus musculus - GenMatch GID: 20068727 | -1..776 | IF | | 100.0 | 57.3 | D |
| pB66_B-6 | 5p/3p | Mus musculus - GenMatch GID: 38093845 | -1..687 | IF | | 100.0 | 85.3 | D |
| pB66_B-20 | 5p/3p | Mus musculus - GenMatch GID: 81295619 | -1 | IF | | ALU 92.7 | 93.8 | D |
| pB66_A-8 | 5p/3p | Mus musculus - GenMatch GID: 25955801 | -1..579 | IF | | 99.8 | 99.8 | D |
| pB66_A-56 | 5p/3p | Mus musculus - GenMatch GID: 39652666 | -1..576 | IF | | 100.0 | 99.8 | D |
| pB66_A-29 | 5p/3p | Mus musculus - GenMatch GID: 39652666 | -1..576 | IF | | 99.8 | 100.0 | D |
| pB66_A-73 | 5p/3p | Mus musculus - GenMatch GID: 53378873 | -1..582 | IF | | 100.0 | 58.3 | D |
| pB66_A-239 | 5p/3p | Mus musculus - GenMatch GID: 21671875 | -1..745 | IF | | 100.0 | 88.9 | D |
| pB66_A-257 | 5p/3p | Mus musculus - GenMatch GID: 84794837 | -1..298 | IF | | 100.0 | 89.5 | D |
| pB66_A-55 | 5p/3p | Mus musculus - GenMatch GID: 37718670 | -1..399 | IF | | 95.2 | 100.0 | D |
| pB66_A-103 | 5p/3p | Mus musculus - GenMatch GID: 27356737 | -1..549 | IF | | 99.8 | 100.0 | D |
| pB66_B-70 | 5p/3p | Mus musculus - GenMatch GID: 51339134 | -1..410 | IF | | 99.0 | 100.0 | D |
| pB66_A-172 | 5p/3p | Mus musculus - GenMatch GID: 51339134 | -1..410 | IF | | 100.0 | 100.0 | D |
| pB66_A-104 | 5p/3p | Mus musculus - GenMatch GID: 58652300 | -1..644 | IF | | 99.1 | 100.0 | D |
| pB66_A-1 | 5p/3p | Mus musculus - GenMatch GID: 76683163 | -1..323 | IF | | 100.0 | 83.1 | D |
| pB66_B-23 | 5p/3p | Mus musculus - GenMatch GID: 26006611 | -1..490 | IF | | 100.0 | 84.7 | D |
| pB66_B-86 | 5p/3p | Mus musculus - GenMatch GID: 47078043 | -1..456 | IF | | 100.0 | 87.5 | D |
| pB66_A-207 | 5p/3p | Mus musculus - GenMatch GID: 47078043 | -1..436 | IF | | 100.0 | 82.5 | D |
| pB66_A-214 | 5p/3p | Mus musculus - GenMatch GID: 58652300 | -1 | IF | | 100.0 | 66.7 | D |
| pB66_A-7 | 5p/3p | Mus musculus - GenMatch GID: 81295619 | -1 | IF | | ALU 92.8 | 93.8 | D |
| pB66_B-87 | 5p/3p | Mus musculus - GenMatch GID: 58332941 | -1..1186 | IF | | 100.0 | 85.5 | D |
| pB66_A-135 | 5p/3p | Mus musculus - GenMatch GID: 33438711 | -1..737 | IF | | 99.7 | 99.4 | D |
| pB66_A-208 | 5p/3p | Mus musculus - GenMatch GID: 33438711 | -1..737 | IF | | 99.0 | 99.1 | D |
| pB66_A-256 | 5p/3p | Mus musculus - GenMatch GID: 33438711 | -1..737 | IF | | 99.7 | 98.8 | D |
| pB66_A-260 | 5p/3p | Mus musculus - GenMatch GID: 33438711 | -1..737 | IF | | 99.7 | 99.2 | D |
| pB66_A-119 | 5p/3p | Mus musculus - GenMatch GID: 33438711 | -1..737 | IF | | 100.0 | 99.7 | D |
| pB66_A-45 | 5p/3p | Mus musculus - GenMatch GID: 33438711 | -1..737 | IF | | 99.9 | 99.2 | D |

| Clone Name | Type Seq | Gene Name (Best Match) | Start..Stop (nt) | Frame | Sens | %Id 5p | %Id 3p | PBS |
|---|---|---|---|---|---|---|---|---|
| pB66_A-86 | 5p/3p | Mus musculus - GenMatch GID: 81295619 | -1 | IF | | ALU 92.8 | 93.8 | D |
| pB66_A-98 | 3p | Mus musculus - GenMatch GID: 50839068 | ..373 | ?? | | | 100.0 | N/A |
| pB66_A-101 | 5p/3p | Mus musculus - GenMatch GID: 81295619 | -1 | IF | | ALU 92.5 | 54.4 | D |
| pB66_B-66 | 5p/3p | Mus musculus - GenMatch GID: 24459826 | -1..393 ✗ | IF | | 100.0 | 87.3 | D |
| pB66_A-59 | 5p/3p | Mus musculus - GenMatch GID: 61741006 | -1..822 ✱ ✗ | IF | | 100.0 | 100.0 | D |
| pB66_A-158 | 5p/3p | Mus musculus - GenMatch GID: 34013588 | -1..1230 ✱ ✗ | IF | | 100.0 | 100.0 | D |
| pB66_A-64 | 5p/3p | Mus musculus - GenMatch GID: 34013588 | -1..1230 ✱ ✗ | IF | | 99.9 | 85.2 | D |
| pB66_A-118 | 5p/3p | Mus musculus - GenMatch GID: 28268663 | -1..790 ✱ ✗ | IF | | 100.0 | 100.0 | D |
| pB66_A-105 | 5p/3p | Mus musculus - GenMatch GID: 72255689 | -1..534 ✱ ✗ | IF | | 100.0 | 100.0 | D |
| pB66_B-74 | 5p/3p | Mus musculus - GenMatch GID: 39725818 | -1..416 ✱ ✗ | IF | | 100.0 | 92.1 | D |
| pB66_A-10 | 5p/3p | Mus musculus - GenMatch GID: 20068621 | -1..880 ✱ ✗ | IF | | 100.0 | 91.8 | D |
| pB66_B-18 | 5p/3p | Mus musculus - GenMatch GID: 45825198 | -1 | IF | | 94.5 | ALU 64.0 | D |
| pB66_B-7 | 5p/3p | Mus musculus - GenMatch GID: 45825198 | -1 | IF | | 85.9 | ALU 64.0 | D |
| pB66_A-49 | 5p/3p | Mus musculus - GenMatch GID: 45825198 | -1 | IF | | 91.8 | ALU 61.3 | D |
| pB66_A-131 | 5p/3p | Mus musculus - GenMatch GID: 45825198 | -1 | IF | | 98.0 | ALU 62.7 | D |
| pB66_B-5 | 5p/3p | Mus musculus - GenMatch GID: 23379824 | -1..537 ✱ ✗ | IF | | 100.0 | 100.0 | D |
| pB66_A-201 | 5p/3p | Mus musculus - GenMatch GID: 23379824 | -1..537 ✱ ✗ | IF | | 99.4 | 100.0 | D |
| pB66_A-184 | 5p/3p | Mus musculus - GenMatch GID: 23379824 | -1..537 ✱ ✗ | IF | | 100.0 | 99.8 | D |
| pB66_A-12 | 5p/3p | Mus musculus - GenMatch GID: 47078043 | -1..462 ✗ | IF | | 100.0 | 84.1 | D |
| pB66_A-15 | 5p/3p | Mus musculus - GenMatch GID: 81295619 | -1 | IF | | ALU 92.9 | 93.8 | D |
| pB66_A-137 | 5p/3p | Mus musculus - GenMatch GID: 73120244 | -1..500 ✱ ✗ | IF | | 100.0 | 100.0 | D |
| pB66_A-16 | 5p/3p | Mus musculus - GenMatch GID: 81295619 | -1 | IF | | ALU 93.1 | 93.8 | D |
| pB66_A-235 | 5p/3p | Mus musculus - GenMatch GID: 20068621 | -1..873 ✱ ✗ | IF | | 96.3 | 99.1 | D |
| pB66_A-267 | 3p | Mus musculus - GenMatch GID: 20068621 | 6..305 ✗ | IF | | | 99.3 | D |
| pB66_A-140 | 5p/3p | Mus musculus - GenMatch GID: 85060584 | -1..275 ✗ | IF | | 99.7 | 65.0 | D |
| pB66_A-139 | 5p/3p | Mus musculus - GenMatch GID: 85060584 | -1..275 ✗ | IF | | 99.1 | 65.0 | D |
| pB66_A-25 | 5p/3p | Mus musculus - GenMatch GID: 81778246 | -1..517 ✱ ✗ | IF | | 100.0 | 92.6 | D |
| pB66_A-236 | 5p/3p | Mus musculus - GenMatch GID: 38564394 | -1..438 ✱ ✗ | IF | | 99.8 | 98.2 | D |
| pB66_A-243 | 5p/3p | Mus musculus - GenMatch GID: 22204310 | -1..850 ✱ ✗ | IF | | 98.7 | 99.9 | D |
| pB66_A-171 | 5p/3p | Mus musculus - GenMatch GID: 81295619 | -1 | IF | | ALU 92.6 | 93.8 | D |

| Clone Name | Type Seq | Gene Name (Best Match) | Start..Stop (nt) | Frame | Sens | %Id 5p | %Id 3p | PBS |
|---|---|---|---|---|---|---|---|---|
| pB66_A-122 | 5p/3p | Mus musculus - GenMatch GID: 76362883 | -1..574 | IF | | 99.8 | 99.8 | D |
| pB66_B-78 | 5p/3p | Mus musculus - GenMatch GID: 76362883 | -1..574 | IF | | 100.0 | 99.7 | D |
| pB66_A-125 | 5p/3p | Mus musculus - GenMatch GID: 76362883 | -1..574 | IF | | 99.1 | 99.8 | D |
| pB66_A-280 | 3p | Mus musculus - GenMatch GID: 76362883 | -1..573 | IF | | | 99.5 | D |
| pB66_A-285 | 5p/3p | Mus musculus - GenMatch GID: 76362883 | -1..574 | IF | | 97.6 | 99.1 | D |
| pB66_A-31 | 5p/3p | Mus musculus - GenMatch GID: 76362883 | -1..574 | IF | | 98.8 | 99.8 | D |
| pB66_A-160 | 5p/3p | Mus musculus - GenMatch GID: 76362883 | -1..574 | IF | | 99.7 | 99.5 | D |
| pB66_A-33 | 5p/3p | Mus musculus - GenMatch GID: 76362883 | -1..574 | IF | | 100.0 | 100.0 | D |
| pB66_A-187 | 5p/3p | Mus musculus - GenMatch GID: 20068621 | -1..874 | IF | | 100.0 | 71.7 | D |
| pB66_A-151 | 5p/3p | Mus musculus - GenMatch GID: 81295619 | -1 | IF | | ALU 92.5 | 93.8 | D |
| pB66_A-156 | 5p/3p | Mus musculus - GenMatch GID: 60459020 | -1..526 | IF | | 100.0 | 88.9 | D |
| pB66_A-68 | 5p/3p | Mus musculus - GenMatch GID: 28881814 | -1..430 | IF | | 98.6 | 99.5 | D |
| pB66_A-77 | 5p/3p | Mus musculus - GenMatch GID: 57012521 | -1..697 | IF | | 99.7 | 100.0 | D |
| pB66_A-79 | 5p/3p | Mus musculus - GenMatch GID: 51315686 | -1..516 | IF | | 94.4 | 100.0 | D |
| pB66_A-109 | 5p/3p | Mus musculus - GenMatch GID: 33285224 | -1..1005 | IF | | 99.8 | 100.0 | D |
| pB66_A-175 | 5p/3p | Mus musculus - GenMatch GID: 33285224 | -1..1005 | IF | | 99.5 | 98.8 | D |
| pB66_A-233 | 5p/3p | Mus musculus - GenMatch GID: 33285224 | -1..1005 | IF | | 99.3 | 99.4 | D |
| pB66_A-136 | 5p/3p | Mus musculus - GenMatch GID: 33285224 | -1..1005 | IF | | 99.9 | 99.7 | D |
| pB66_A-124 | 5p/3p | Mus musculus - GenMatch GID: 33285224 | -1..1005 | IF | | 100.0 | 99.5 | D |
| pB66_A-189 | 5p/3p | Mus musculus - GenMatch GID: 12642989 | -1 | IF | | 100.0 | 60.6 | D |
| pB66_A-145 | 5p/3p | Mus musculus - GenMatch GID: 24527497 | -1..383 | IF | | 99.2 | 97.4 | D |
| pB66_B-2 | 5p/3p | Mus musculus - GenMatch GID: 42475643 | -1..405 | IF | | 99.8 | 100.0 | D |
| pB66_A-14 | 5p/3p | Mus musculus - GenMatch GID: 28300643 | -1..313 | IF | | 100.0 | 100.0 | D |
| pB66_A-199 | 5p/3p | Mus musculus - GenMatch GID: 34787446 | -1..509 | IF | | 100.0 | 89.8 | D |
| pB66_B-57 | 5p/3p | Mus musculus - GenMatch GID: 21322392 | -1..501 | IF | | 98.4 | 100.0 | D |
| pB66_A-244 | 5p/3p | Mus musculus - GenMatch GID: 24459826 | -1..393 | IF | | 100.0 | 100.0 | D |
| pB66_A-193 | 5p/3p | Mus musculus - GenMatch GID: 24459826 | -1..393 | IF | | 100.0 | 99.5 | D |
| pB66_B-59 | 5p/3p | Mus musculus - GenMatch GID: 24459826 | -1..393 | IF | | 100.0 | 99.7 | D |
| pB66_A-163 | 5p/3p | Mus musculus - GenMatch GID: 24459826 | -1..393 | IF | | 100.0 | 100.0 | D |
| pB66_A-23 | 5p/3p | Mus musculus - GenMatch GID: 24459826 | -1..393 | IF | | 100.0 | 100.0 | D |

| Clone Name | Type Seq | Gene Name (Best Match) | Start..Stop (nt) | Frame | Sens | %Id 5p | %Id 3p | PBS |
|---|---|---|---|---|---|---|---|---|
| pB66_A-170 | 5p/3p | Mus musculus - GenMatch GID: 24459826 | -1..393 | IF | | 100.0 | 100.0 | D |
| pB66_A-144 | 5p/3p | Mus musculus - GenMatch GID: 24459826 | -1..393 | IF | | 100.0 | 100.0 | D |
| pB66_A-192 | 5p/3p | Mus musculus - GenMatch GID: 24459826 | -1..393 | IF | | 100.0 | 100.0 | D |
| pB66_A-95 | 5p/3p | Mus musculus - GenMatch GID: 24459826 | -1..393 | IF | | 100.0 | 100.0 | D |
| pB66_A-19 | 5p/3p | Mus musculus - GenMatch GID: 24459826 | -1..393 | IF | | 100.0 | 100.0 | D |
| pB66_A-2 | 5p/3p | Mus musculus - GenMatch GID: 24459826 | -1..393 | IF | | 100.0 | 100.0 | D |
| pB66_A-264 | 5p/3p | Mus musculus - GenMatch GID: 81295619 | -1 | IF | | ALU 92.5 | 93.8 | D |
| pB66_A-271 | 5p | Mus musculus - GenMatch GID: 34787446 | -1 | IF | | 100.0 | | D |
| pB66_A-274 | 5p/3p | Mus musculus - GenMatch GID: 49035075 | -1..780 | IF | | 100.0 | 95.1 | D |
| pB66_A-147 | 5p/3p | Mus musculus - GenMatch GID: 74039307 | -1..682 | IF | | 99.4 | 99.7 | D |
| pB66_B-27 | 3p | Mus musculus - GenMatch GID: 21065354 | ..666 | ?? | | | 100.0 | N/A |
| pB66_A-246 | 3p | Mus musculus - GenMatch GID: 47078043 | -1..463 | IF | | | 100.0 | D |
| pB66_A-220 | 5p/3p | Mus musculus - GenMatch GID: 47078043 | -1..463 | IF | | 97.4 | 99.8 | D |
| pB66_A-58 | 5p/3p | Mus musculus - GenMatch GID: 47078043 | -1..463 | IF | | 100.0 | 100.0 | D |
| pB66_A-67 | 5p/3p | Mus musculus - GenMatch GID: 47078043 | -1..463 | IF | | 100.0 | 100.0 | D |
| pB66_A-224 | 5p/3p | Mus musculus - GenMatch GID: 47078043 | -1..463 | IF | | 100.0 | 100.0 | D |
| pB66_A-44 | 5p/3p | Mus musculus - GenMatch GID: 47078043 | -1..463 | IF | | 96.6 | 100.0 | D |
| pB66_B-65 | 3p | Mus musculus - GenMatch GID: 47078043 | ..539 | ?? | | | 99.0 | N/A |
| pB66_A-275 | 5p | Mus musculus - GenMatch GID: 58652300 | -1 | IF | | 100.0 | | D |
| pB66_B-63 | 5p/3p | Mus musculus - GenMatch GID: 62123204 | -1..1255 | IF | | 100.0 | 93.6 | C |
| pB66_A-81 | 5p/3p | Mus musculus - GenMatch GID: 62123204 | 9..424 | IF | | 100.0 | 97.1 | C |
| pB66_A-130 | 5p/3p | Mus musculus - GenMatch GID: 62123204 | 9..424 | IF | | 100.0 | 99.5 | C |
| pB66_B-13 | 5p/3p | Mus musculus - GenMatch GID: 34495144 | -1 | IF | | 98.5 | 83.3 | D |
| pB66_A-133 | 5p/3p | Mus musculus - GenMatch GID: 34495144 | -1 | IF | | 99.6 | ALU 83.3 | D |
| pB66_A-82 | 5p/3p | Mus musculus - GenMatch GID: 34495144 | -1 | IF | | 98.6 | 83.3 | D |
| pB66_B-49 | 5p/3p | Mus musculus - GenMatch GID: 33438668 | -1..589 | OOF1 | | 100.0 | 86.6 | N/A |
| pB66_B-48 | 5p/3p | Mus musculus - GenMatch GID: 21671875 | -1..744 | IF | | 98.1 | 98.6 | D |
| pB66_B-15 | 5p/3p | Mus musculus - GenMatch GID: 21671875 | -1..744 | IF | | 97.7 | 99.2 | D |
| pB66_A-48 | 5p/3p | Mus musculus - GenMatch GID: 21671875 | -1..744 | IF | | 97.2 | 99.2 | D |
| pB66_A-240 | 5p/3p | Mus musculus - GenMatch GID: 21671875 | -1..738 | IF | | 100.0 | 82.2 | D |

| | | | | | | | | | |
|---|---|---|---|---|---|---|---|---|---|
| pB66_A-186 | 5p/3p | Mus musculus - GenMatch GID: 26006611 | -1..511 | ✕ | IF | | 100.0 | 100.0 | D |
| pB66_A-153 | 5p/3p | Mus musculus - GenMatch GID: 26006611 | -1..511 | ✕ | IF | | 99.8 | 99.8 | D |
| pB66_B-31 | 5p/3p | Mus musculus - GenMatch GID: 37719149 | -1..343 | ✻ ✕ | IF | | 100.0 | 82.6 | D |
| pB66_B-92 | 5p/3p | Mus musculus - GenMatch GID: 30985141 | -1..489 | ✕ | IF | | 100.0 | 95.5 | D |
| pB66_A-100 | 5p/3p | Mus musculus - GenMatch GID: 30985141 | -1..490 | ✻ ✕ | IF | | 100.0 | 87.6 | D |
| pB66_A-121 | 5p/3p | Mus musculus - GenMatch GID: 68270833 | -1..386 | ✻ ✕ | IF | | 100.0 | 100.0 | D |
| pB66_B-81 | 5p/3p | Mus musculus - GenMatch GID: 82465620 | -1..565 | ✕ | IF | | 100.0 | 60.6 | D |
| pB66_A-20 | 5p/3p | Mus musculus - GenMatch GID: 66773599 | -1..783 | ✻ ✕ | IF | | 99.7 | 99.8 | D |
| pB66_B-68 | 5p/3p | Mus musculus - GenMatch GID: 76362883 | -1..571 | ✕ | IF | | 100.0 | 93.2 | D |
| pB66_B-67 | 5p/3p | Mus musculus - GenMatch GID: 33438668 | -1..624 | ✻ ✕ | OOF1 | | 98.7 | 93.1 | N/A |
| pB66_A-277 | 5p/3p | Mus musculus - GenMatch GID: 50839068 | -1 | | IF | | 96.1 | 78.6 | D |
| pB66_B-32 | 5p/3p | Mus musculus - GenMatch GID: 50839068 | -1 | | IF | | 98.7 | 77.8 | D |
| pB66_A-123 | 5p/3p | Mus musculus - GenMatch GID: 50839068 | -1 | | IF | | 99.9 | 68.2 | D |
| pB66_A-129 | 5p/3p | Mus musculus - GenMatch GID: 50839068 | -1 | | IF | | 99.8 | 68.2 | D |
| pB66_B-14 | 5p/3p | Mus musculus - GenMatch GID: 81778322 | -1..1165 | ✻ ✕ | OOF1 | | 100.0 | 93.6 | N/A |
| pB66_A-111 | 5p/3p | Mus musculus - GenMatch GID: 81778246 | -1..517 | ✕ | IF | | 98.5 | 99.8 | D |
| pB66_A-66 | 5p/3p | Mus musculus - GenMatch GID: 60459020 | -1..533 | ✻ ✕ | IF | | 99.8 | 100.0 | D |
| pB66_A-62 | 5p/3p | Mus musculus - GenMatch GID: 60459020 | -1..533 | ✻ ✕ | IF | | 100.0 | 100.0 | D |
| pB66_B-9 | 3p | Mus musculus - GenMatch GID: 60459020 | ..1047 | | ?? | | | 100.0 | D |
| pB66_A-197 | 3p | Mus musculus - GenMatch GID: 60459020 | ..490 | | ?? | | | 99.2 | D |
| pB66_A-219 | 5p/3p | Mus musculus - GenMatch GID: 76683163 | -1..323 | ✕ | IF | | 99.4 | 94.1 | D |
| pB66_A-206 | 5p/3p | Mus musculus - GenMatch GID: 76683163 | -1..323 | ✕ | IF | | 98.5 | 94.1 | D |
| pB66_A-204 | 5p/3p | Mus musculus - GenMatch GID: 76683163 | -1..323 | ✕ | IF | | 98.8 | 94.4 | D |
| pB66_A-36 | 5p/3p | Mus musculus - GenMatch GID: 81295619 | -1 | | IF | | ALU 92.9 | 93.8 | D |
| pB66_A-141 | 5p/3p | Mus musculus - GenMatch GID: 84794837 | -1..314 | ✕ | IF | | 100.0 | 100.0 | D |
| pB66_A-6 | 5p/3p | Mus musculus - GenMatch GID: 84794837 | -1..314 | ✕ | IF | | 100.0 | 100.0 | D |
| pB66_A-39 | 3p | Mus musculus - GenMatch GID: 34495144 | ..645 | | ?? | | | 97.5 | N/A |
| pB66_A-113 | 5p/3p | Mus musculus - GenMatch GID: 83313863 | -1 | | IF | | 100.0 | 64.7 | D |
| pB66_A-200 | 5p/3p | Mus musculus - GenMatch GID: 83313863 | -1 | | IF | | 99.7 | 64.7 | D |
| pB66_A-43 | 5p/3p | Mus musculus - GenMatch GID: 83313863 | -1 | | IF | | 100.0 | 64.7 | D |

HYBRiGENiCS

services

| | | | | | | | | | |
|---|---|---|---|---|---|---|---|---|---|
| pB66_A-72 | 5p/3p | Mus musculus - GenMatch GID: 83313863 | -1 | | IF | | 99.8 | 64.7 | D |
| pB66_A-230 | 5p/3p | Mus musculus - GenMatch GID: 81295619 | -1 | | IF | | 100.0 | 65.1 | D |
| pB66_B-50 | 5p/3p | 81295619 Mus musculus - GenMatch GID: | -1 | | IF | | ALU 92.8 | 93.8 | D |
| pB66_A-63 | 5p/3p | Mus musculus - GenMatch GID: 45825198 | -1..609 | ✗ | IF | | 100.0 | ALU 60.2 | D |
