## Supplementary figures and images for "Identification of novel microcephaly-linked protein ABBA that mediates cortical progenitor cell division and corticogenesis through NEDD9-RhoA"

### supplemental figure 1

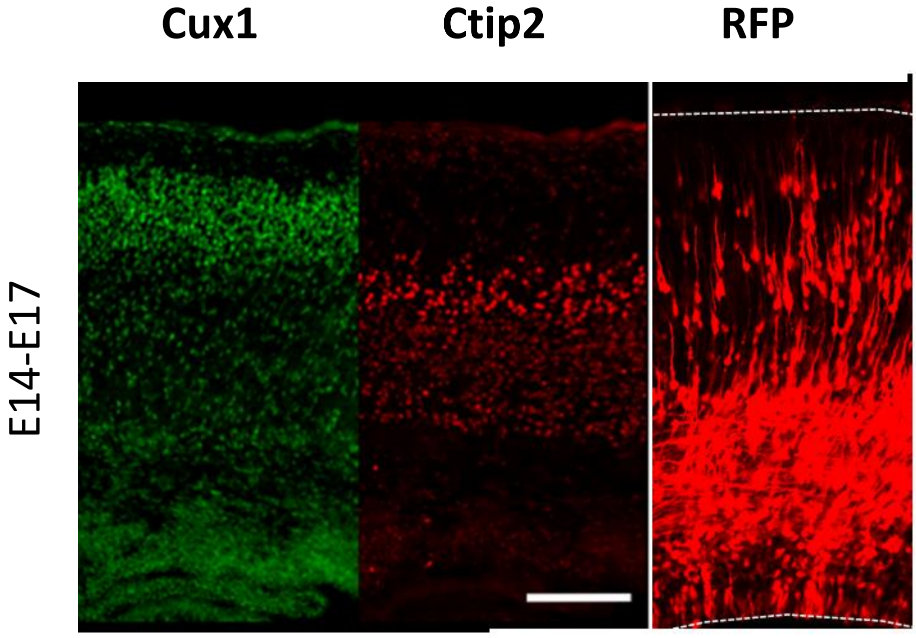

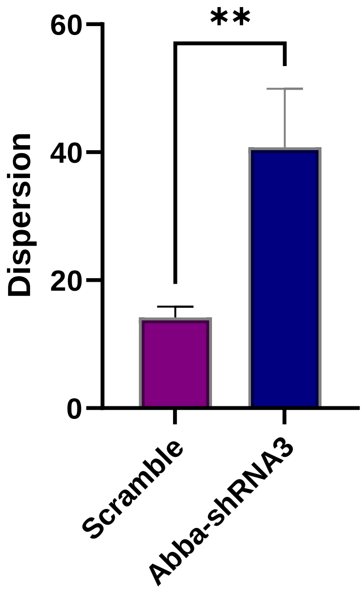

### supplemental figure 2

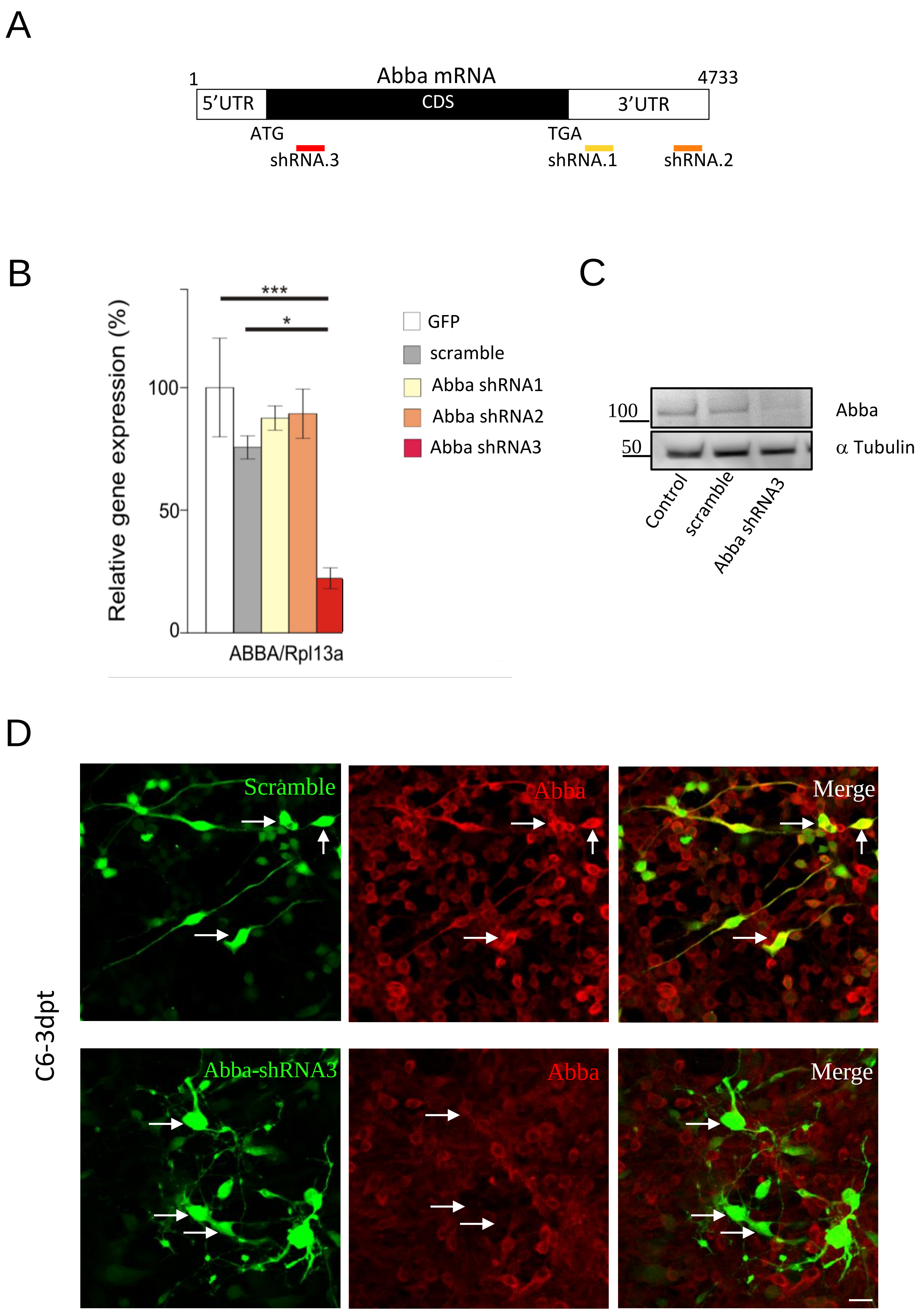

### supplemental figure 3

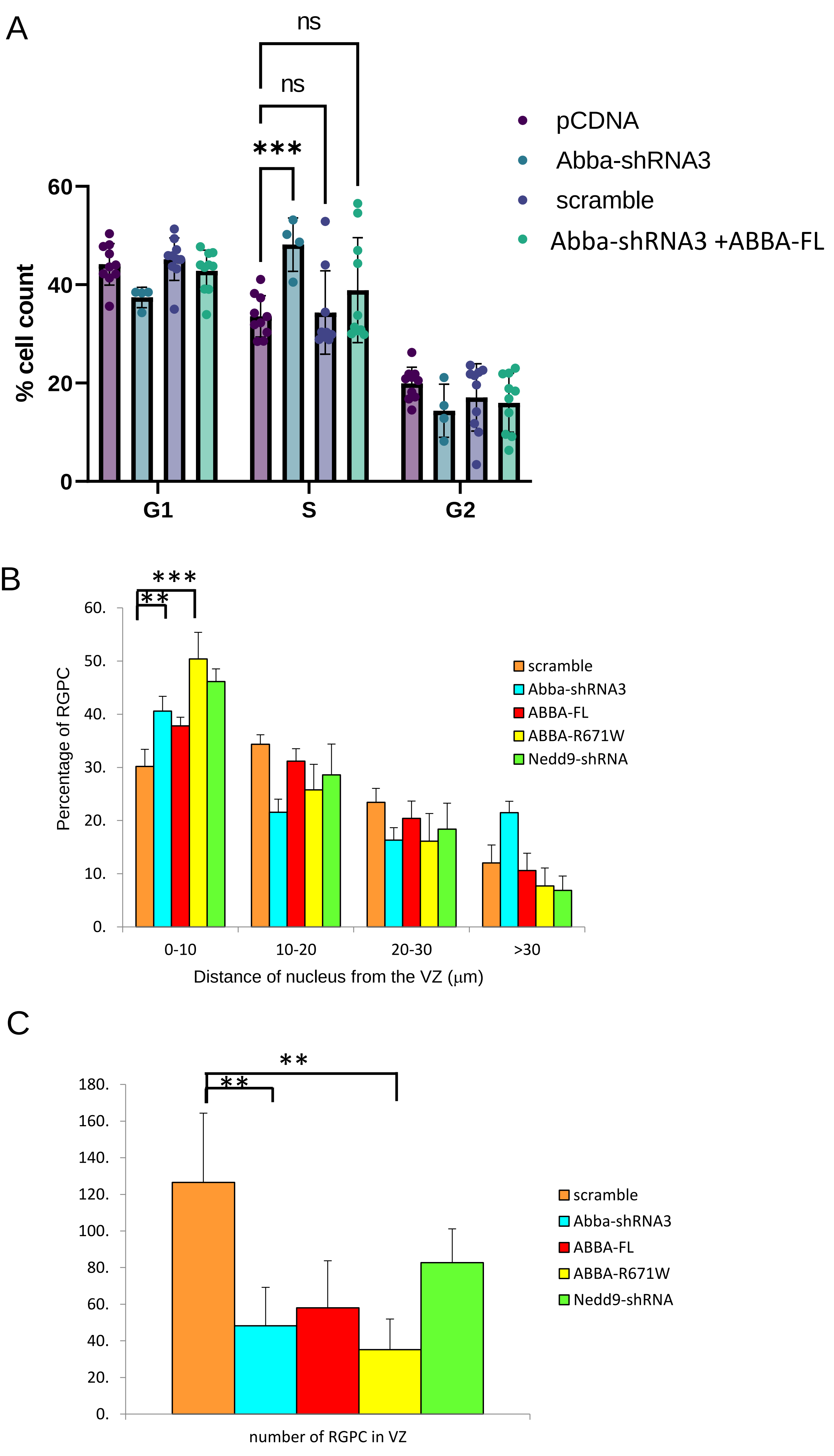

### supplemental figure 4

Sup fig 4

A

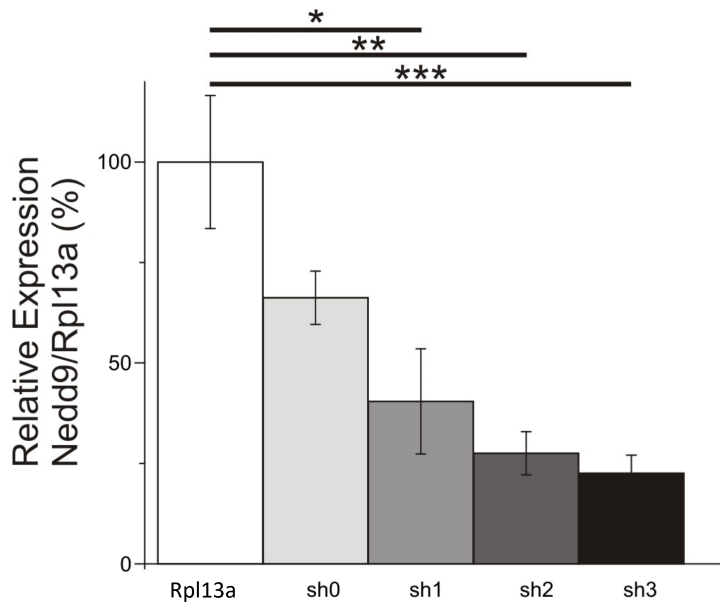


B

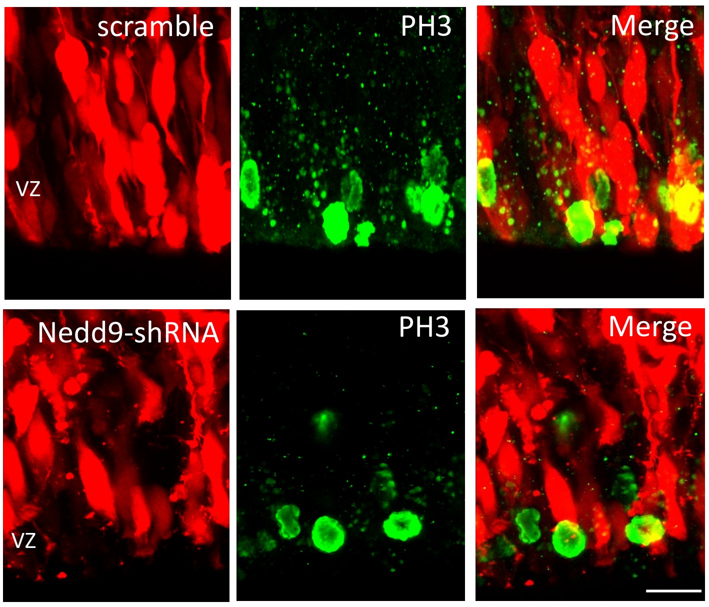


C

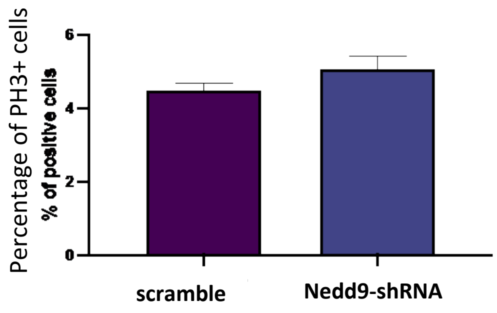


D

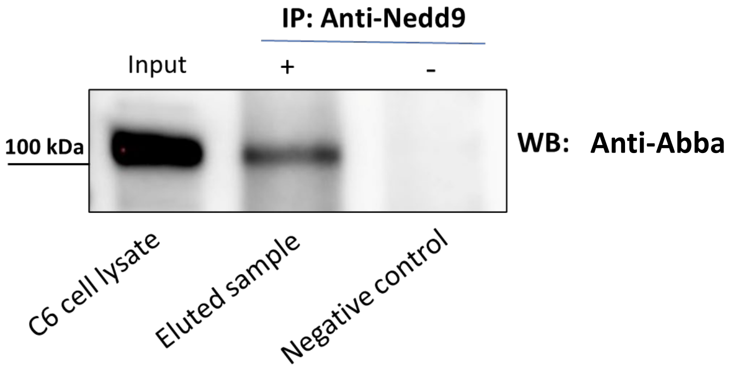

### supplemental figure 5

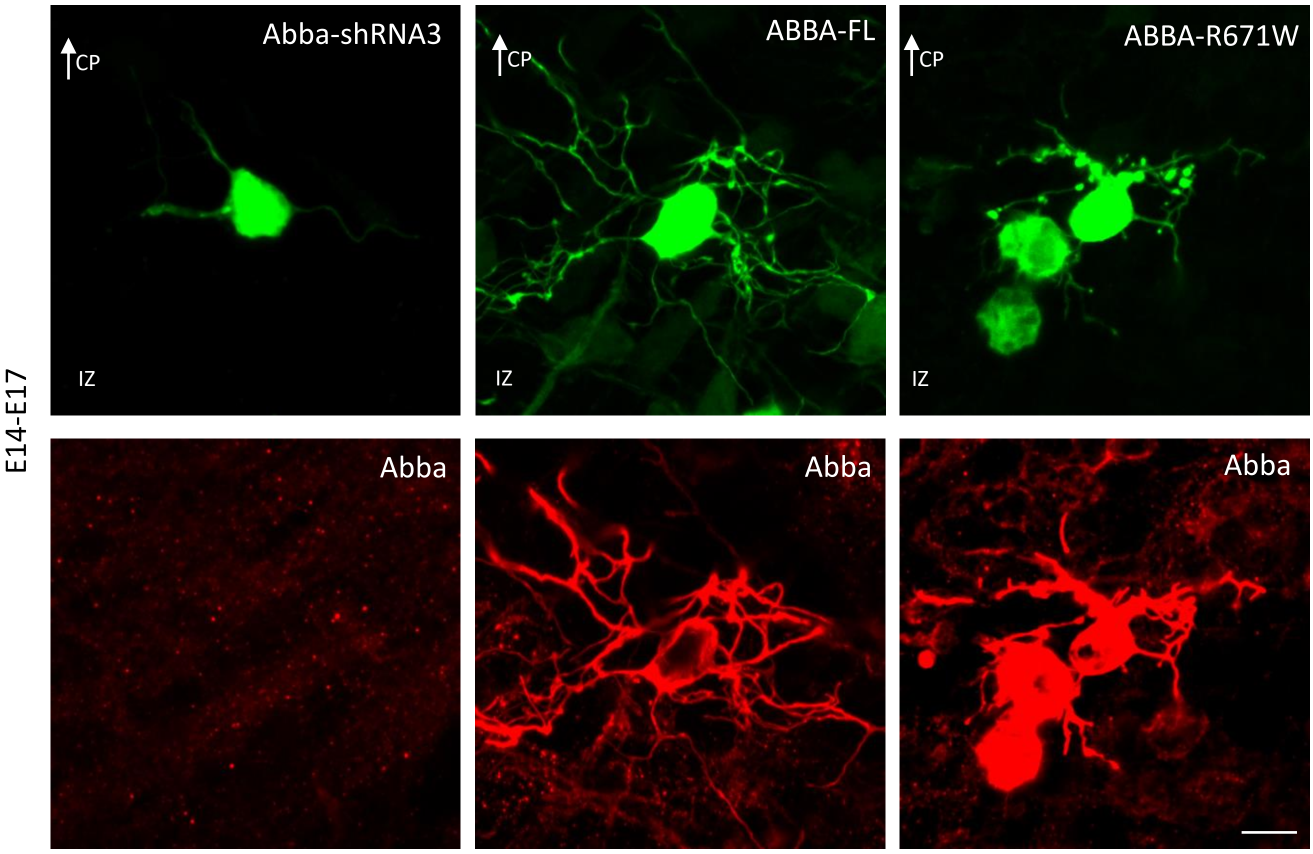

### supplemental figure 6

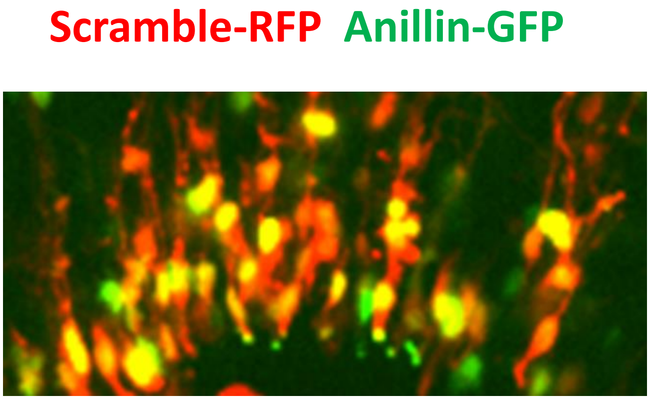

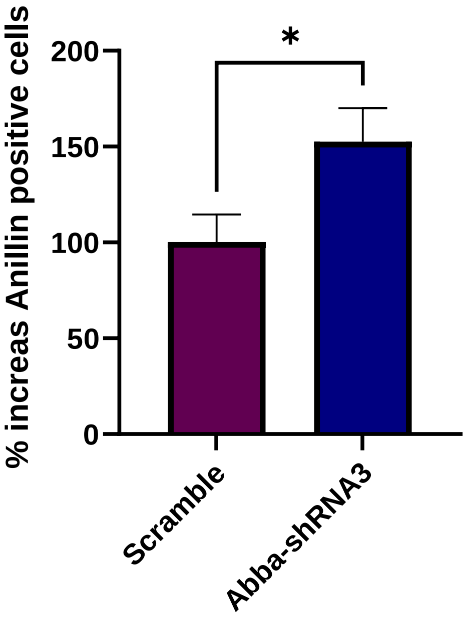
