## Supplementary material for "Identification of novel microcephaly-linked protein ABBA that mediates cortical progenitor cell division and corticogenesis through NEDD9-RhoA": table 1

| | |
|---|---|
| Variant | c.2011C>T, heterozygous |
| | p.R671W |
| | *de novo* |
| age | In their 10's |
| Gender | M |
| Family History | Caucasian, dutch |
| | 27 |
| Growth | (- 2SD) |
| Microcephaly | Borderline microcephaly |
| Neurological examination | yes |
| Intellectual disability | Yes, mild ID IQ69 |
| Seizures | No |
| Behavioral problems | Autism spectrum disorder, ADHD |
| Ophthalmological anomalies | Congenital nystagmus |
| Craniofacial dysmorphisms | Small ears with abnormal hlices, ptosis left, epicanthus, diastasis teeth, |
| Urorenal abnormalities | No |
| Skeletal abnormalities | No |
